# Long-read sequencing reveals the RNA isoform repertoire of neuropsychiatric risk genes in human brain

**DOI:** 10.1101/2024.02.22.24303189

**Authors:** Ricardo De Paoli-Iseppi, Shweta Joshi, Josie Gleeson, Yair David Joseph Prawer, Yupei You, Ria Agarwal, Anran Li, Anthea Hull, Eloise Marie Whitehead, Yoonji Seo, Rhea Kujawa, Raphael Chang, Mriga Dutt, Catriona McLean, Benjamin Leo Parker, Michael Ben Clark

## Abstract

Neuropsychiatric disorders are highly complex conditions and the risk of developing a disorder has been tied to hundreds of genomic variants that alter the expression and/or products (isoforms) made by risk genes. However, how these genes contribute to disease risk and onset through altered expression and RNA splicing is not well understood. Combining our new bioinformatic pipeline IsoLamp with nanopore long-read amplicon sequencing, we deeply profiled the RNA isoform repertoire of 31 high-confidence neuropsychiatric disorder risk genes in human brain. We show most risk genes are more complex than previously reported, identifying 363 novel isoforms and 28 novel exons, including isoforms which alter protein domains, and genes such as *ATG13* and *GATAD2A* where most expression was from previously undiscovered isoforms. The greatest isoform diversity was present in the schizophrenia risk gene *ITIH4*. Mass spectrometry of brain protein isolates confirmed translation of a novel exon skipping event in ITIH4, suggesting a new regulatory mechanism for this gene in brain. Our results emphasize the widespread presence of previously undetected RNA and protein isoforms in brain and provide an effective approach to address this knowledge gap. Uncovering the isoform repertoire of neuropsychiatric risk genes will underpin future analyses of the functional impact these isoforms have on neuropsychiatric disorders, enabling the translation of genomic findings into a pathophysiological understanding of disease.

## Introduction

Over 90% of multi-exonic human genes undergo alternative splicing (AS), a process that enables genes to produce multiple mRNA products (RNA isoforms) [1]. Common AS events include exon skipping, intron retention and alternative 5’ and 3’ exonic splice sites [2]. These mRNA alterations can impact the open reading frame (ORF) and/or alter post-transcription regulation of an RNA, increasing both transcriptomic and proteomic diversity [1, 3, 4]. AS has been established as an important regulator of organ development and physiological functions and is highly regulated under normal conditions [5, 6]. Conversely, aberrant RNA splicing has been linked to the development of cancer, autoimmune and neurodevelopmental disorders [7–11]. AS plays an especially important role in the brain, which has a distinct splicing program including the largest number of tissue specific exons and frequent use of microexons [12]. Numerous studies have reported crucial roles for AS in brain development and dysregulation in disease [13, 14].

Neuropsychiatric or mental health disorders (MHDs) including schizophrenia (SZ), major depressive disorder (MDD), autism spectrum disorder (ASD) and bipolar disorder (BD) can carry significant morbidity for affected individuals [15]. Comorbidities, delayed diagnoses and stigma surrounding MHDs also present a significant challenge to individuals and their families [16]. However, treatment options remain limited or are not well tolerated or effective in some individuals and the underlying aetiology of disease and risk remains poorly understood [17–19]. Recently, genome wide association studies (GWAS) have revealed hundreds of common single nucleotide polymorphisms (SNPs) that are associated with risk of developing neuropsychiatric disease [20–25]. Confirmatory studies including transcriptome wide association studies (TWAS), summary data–based Mendelian randomization (SMR) [26], multimarker analysis of genomic annotation (MAGMA) and variants (H-MAGMA [27], nMAGMA [28]) and functional genomics have helped to identify the risk genes at these loci and also showed a considerable number of risk loci are shared between disorders [29].

Understanding how risk variants affect risk genes is not straightforward, the vast majority of risk variants are found in non-coding parts of the genome and are expected to be regulatory, impacting gene expression levels or which RNA isoforms are produced. Risk variants may impact splicing factor binding leading to altered isoform splicing ratios [8]. For example, a risk variant block (rs1006737) within intron 3 of the SZ risk gene *CACNA1C* was linked to variable mRNA expression, while *GAD1* long and short isoform expression in hippocampus was associated with the SZ and ASD risk variant (rs3749034) within the promoter [30, 31]. However, there is a current lack of understanding about how risk gene expression and splicing are altered by the risk variants and therefore profiling both their expression and RNA isoforms is essential to link genetic changes to disease pathophysiology.

Current sequencing technologies including Illumina short-reads perform well at detecting novel AS, however the lack of long-range exon connectivity information inherent in short-reads means these approaches are limited in their ability to identify and quantify full-length isoforms and this issue is exacerbated in longer, more complex genes [32, 33]. In contrast, long-read technologies including Oxford Nanopore Technologies (ONT) and Pacific Biosciences (PacBio) can sequence entire isoforms in a single read enabling more accurate isoform profiling [7, 34]. Such technologies now make it feasible to comprehensively examine gene isoform profiles. Initial investigations of *SNX19* and *CACNA1C* demonstrated the incomplete knowledge of isoform profiles in humans and the likely importance of novel gene isoforms in disease risk [35, 36].

In this study we addressed the lack of knowledge surrounding MHD risk gene isoform expression using nanopore amplicon sequencing. We developed a new bioinformatic tool, IsoLamp, to identify known and novel RNA isoforms from long-read data. Analysis of the RNA splicing profiles of 31 MHD risk genes identified 363 novel RNA isoforms and 28 novel exons. We identified several genes where most expression is from novel isoforms, including *ATG13* and *GATAD2A*, where the most highly expressed isoforms were novel. Our results show the transcript structure for most risk genes is more complex than current annotations, containing additional exon skipping events, retained introns, novel splice sites and novel exons, including novel isoforms that alter the protein and potentially its function. This work lays the foundation for a better understanding of how risk gene isoforms may play a role in disease pathophysiology.

## Results

### Experimental overview

To identify the RNA isoforms expressed from genes of interest, collated from GWAS evidence, we aimed to perform long-read amplicon sequencing, which provides a highly sensitive means for comprehensive isoform discovery and relative quantification (Figure 1A, Supplementary Figure 1) [35]. We selected seven regions of post-mortem human brain from five control individuals, encompassing both transcriptionally divergent regions as well as those highly implicated in MHDs (Supplementary Table 1). Amplicons were designed cover the full coding region of target genes and, where possible, run from the first to the last exon. Multiple set of primers were used for genes with alternative transcriptional initiation and termination exons and/or alternative coding sequence initiation and termination sites.

**Figure 1.**
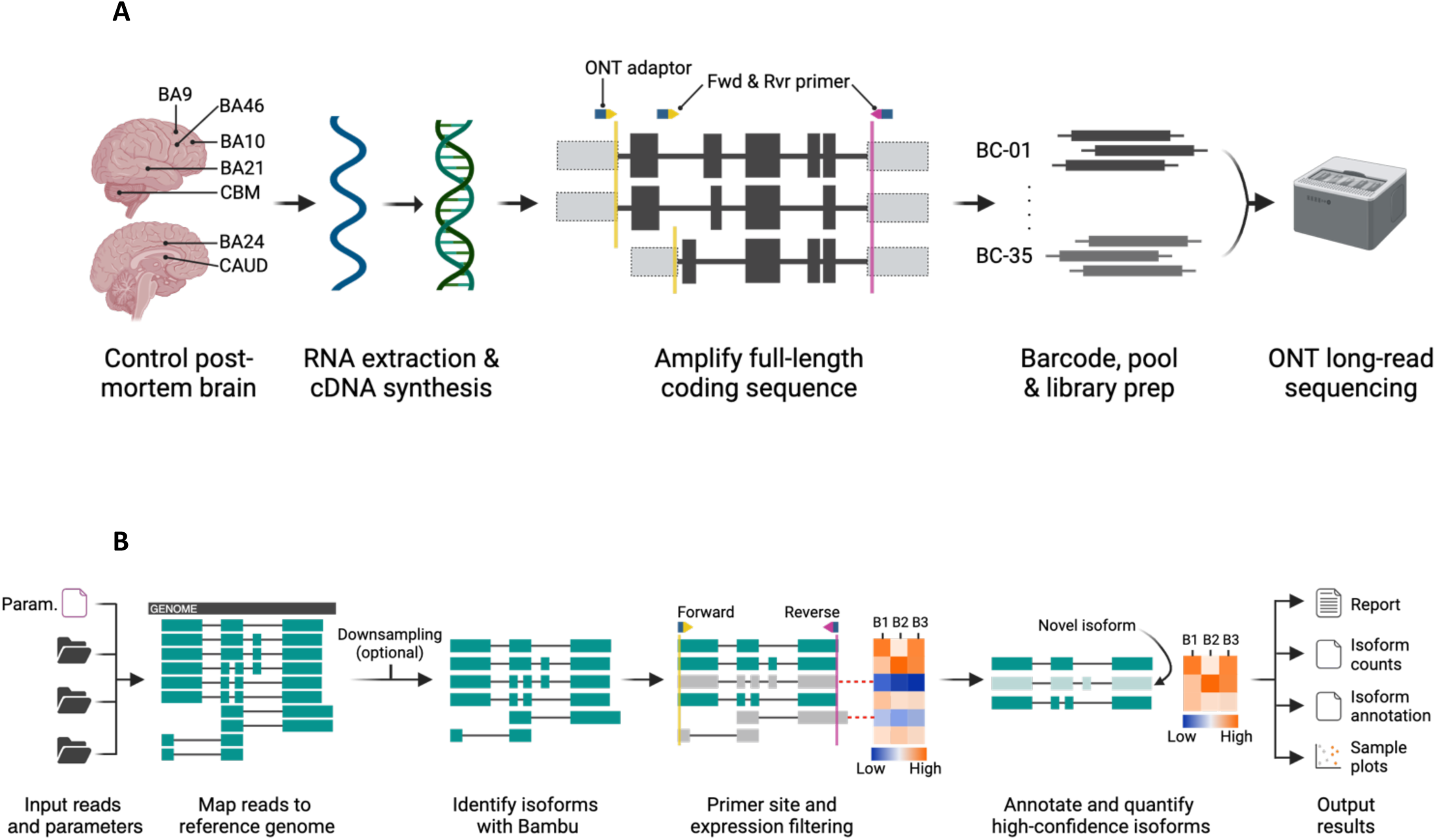
**A. RNA isoform sequencing of human post-mortem brain.** RNA was isolated from frontal cortical regions, caudate (CAUD) and cerebellum (CBM) and converted to cDNA. The coding sequence (black boxes) was amplified using specific forward (Fwd, yellow arrows) and reverse (Rvr, pink arrow) primers generally designed in the 5’ and 3’ UTR regions (grey boxes) to capture as many isoforms as possible. An Oxford Nanopore Technologies (ONT) adaptor sequence (blue box) was incorporated into each primer for sample multiplexing. Samples were then barcoded and pooled to create a single library for long-read sequencing on a GridION. Key: Brodmann Area (BA), barcode (BC), Oxford Nanopore Technologies (ONT). **B. Isoform discovery with long-read amplicon sequencing (IsoLamp) workflow.** A gene specific parameters file (containing chromosome and primer coordinates) was used to align long-reads for each sample (B1-3) against the reference (black box) using Minimap2. Known and novel RNA isoforms were identified using Bambu. Identified isoforms are then filtered (grey isoforms) to remove those not overlapping forward (yellow line) and reverse (pink line) primer positions, ensuring full-length isoform discovery. Low expression (blue on heatmap) isoforms were also filtered out as indicated by dashed red lines. Filtered known and novel isoforms can then be annotated, quantified using IsoLamp output files and visualised using IsoVis.

### IsoLamp: a tool for RNA isoform discovery from long-read amplicon sequencing

While there are several long-read isoform discovery and quantification tools, these are not generally optimised for amplicon sequencing of single genes at high depth. Therefore, we created ISOform discovery with Long-read AMPlicon sequencing (IsoLamp), a custom pipeline designed for isoform profiling from amplicon sequencing (Figure 1B). In contrast to previous tools [35] IsoLamp can be applied to any gene, provides flexible filtering options and provides a simpler, unified, output of isoforms.

We benchmarked the performance of IsoLamp using synthetic Spike-in RNA variants (SIRVs) that provide a known ground truth for isoform exonic structures and abundances. We performed long-read amplicon sequencing on SIRV5 and SIRV6, targeting five isoforms per gene, as these SIRVs allowed targeting of the largest number of isoforms with a single primer pair and so best recapitulated human genes. The SIRV dataset comprised nine replicates from each of the three SIRV mixes (E0, E1, E2) for each gene (Supplementary Figure 2). 99% of reads mapped to the SIRV genome with minimap2 [37], confirming on-target amplification. We benchmarked the performance of IsoLamp with Bambu [38], FLAIR [39], FLAMES [40] and Stringtie2 (-L) [41]. We assessed the precision, recall and expression correlations of the five tools using three different reference annotations (Figure 2A-C, Supplementary Figure 3, Supplementary Table 2): **1.** Complete - contains all SIRV isoforms, **2.** Insufficient - missing SIRV isoforms known to be present, and **3.** Over – contains additional isoforms that are not present in the SIRV mixes.

**Figure 2.**
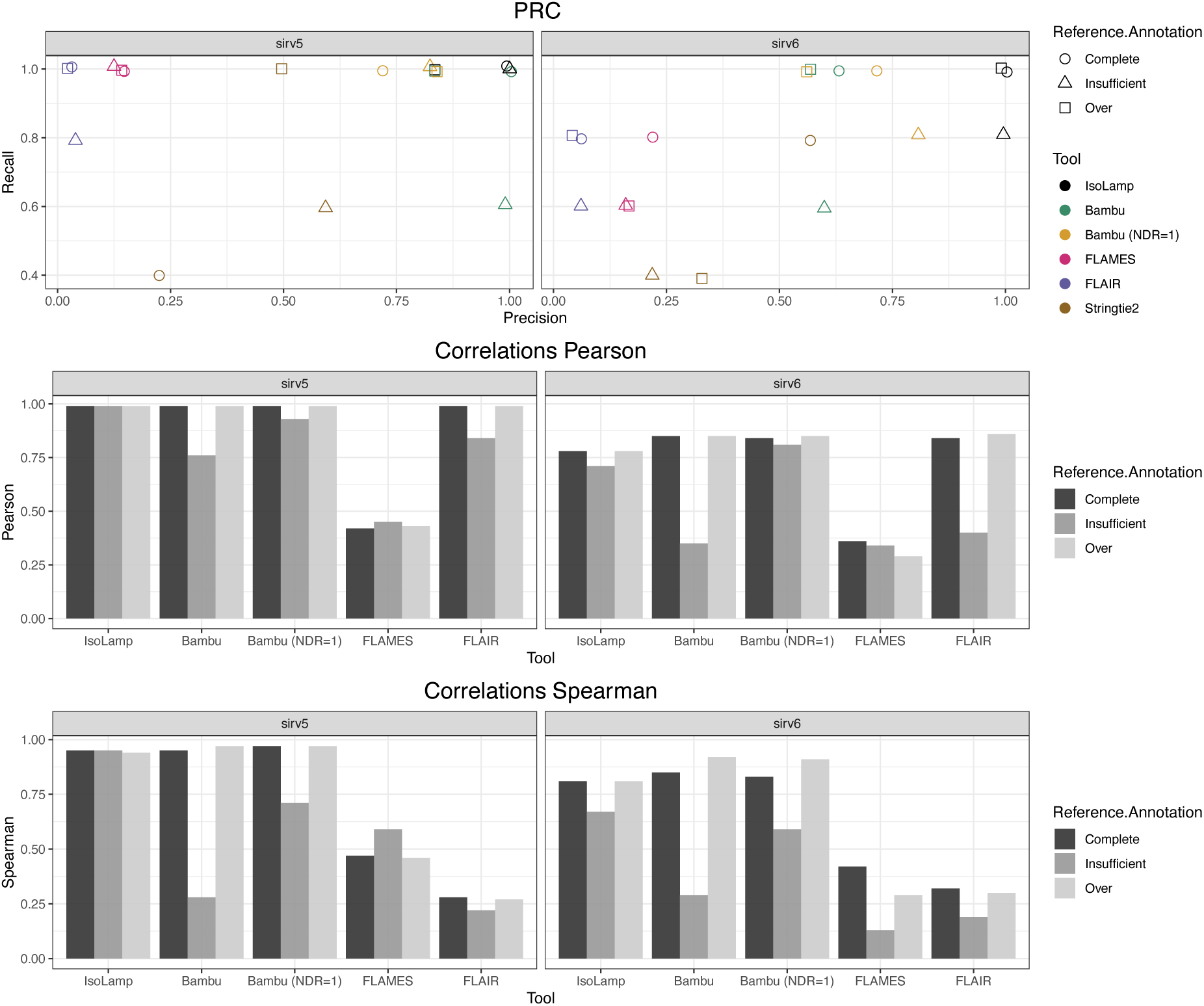
Benchmarking IsoLamp using spike-in SIRVs. **A.** Precision recall of each tested pipeline with the complete, insufficient or over annotated SIRV reference. IsoLamp (black) returned high quality isoforms from amplicon data of both SIRV5 and 6. Pearson **(B)** and Spearman **(C)** correlations for each pipeline between known and observed expression values for SIRV 5 and 6 mixes.

Our benchmarking results demonstrated IsoLamp had the highest precision and recall values, consistently outperforming other isoform discovery tools by correctly identifying true isoforms and minimising false positives (Figure 2A, Supplementary Table 2). This included maintaining high performance with the more challenging, but also more realistic, insufficient and over annotation references. IsoLamp expression quantification was also consistently accurate and maintained performance irrespective of the annotation provided (Figure 2B, C).

Bambu, which is also utilised within the IsoLamp pipeline, was the next best performing tool, although it identified more false positives and had poorer recall and quantification results using the insufficient annotation (Figure 2A-C). IsoLamp utilises Bambu parameters optimised for amplicon-sequencing, including a novel discovery rate (NDR) of 1. Adjusting the Bambu ‘NDR’ to 1 improved its recall but didn’t improve precision (Figure 2A-C, Supplementary Table 2). These results demonstrate how IsoLamp outperforms tools designed for whole-transcriptome analysis, including when Bambu is provided with optimised isoform discovery parameters for amplicon sequencing.

FLAIR had the highest number of isoforms of all tools tested identifying 261, 181, and 278 novel transcripts in the complete, insufficient, and over-annotated reference-based analyses, respectively. This high level of false-positive novel transcripts led to inaccuracies in transcript abundance assignments, resulting in low correlations compared to other tools (Figure 2A-C). FLAMES exhibited 100% recall for SIRV5 across all annotations, however, its performance with SIRV6 was suboptimal, indicating a higher degree of variability in the FLAMES isoform discovery pipeline. FLAMES also performed poorly for isoform quantification. Lastly, while Stringtie2 did not introduce large numbers of false positives, it had the highest number of false negatives, including when provided with complete annotations (Figure 2A-C, Supplementary Table 2).

IsoLamp employs an optimised expression-based filter to remove lowly expressed isoforms that are likely to be false positive detections. Applying this filter to Bambu, FLAIR, and FLAMES substantially reduced false positive novel isoforms and enhanced overall precision (Supplementary Figure 3, Supplementary Table 2), though IsoLamp was still the top performing tool. Beyond synthetic benchmarking data, reference annotations are typically a combination of insufficient and over annotations. In such scenarios, IsoLamp demonstrated better or comparable correlations with all other tools (Figure 1A-C, Supplementary Figure 3), suggesting its superiority for amplicon-sequencing based isoform discovery and quantification from real biological data.

### Post-mortem human brain RNA quantity and quality

Total RNA for long-range amplicon sequencing was extracted from 7 brain regions from 5 healthy individuals (Ind01 - 05) and subject to sample QC (Supplementary Figure 4A-D). RINe (mean = 7.4, range = 6 - 8.1) did not differ by brain region, however Ind04 had significantly lower RINe scores (Supplementary Figure 4B). No trend between the PMI (mean = 44.25 hrs) and RINe was observed (Supplementary Figure 4C). RINe appeared to worsen with decreasing brain tissue pH levels (Supplementary Figure 4D). A principal component analysis (PCA) showed separation of Ind04 (likely driven by lower sample pH and RINe) and of cerebellum and caudate samples from cortical regions in PC1 and PC2 (Supplementary Figure 5AB). A relatively small proportion of variance (4.7%) was attributed to control donor age in PC6 (Supplementary Figure 5C).

### Long read sequencing identifies 363 novel RNA isoforms

A total of 31 risk genes were selected for amplicon sequencing based on the accumulated evidence for their involvement in neuropsychiatric disorder risk. A custom database of risk genes and their evidence levels was created and genes ranked (Methods). In a reflection of current GWAS cohort sizes, 21 of the selected genes had the highest evidence for involvement in risk for SZ, 7 for MDD, 2 for ASD and 1 for BPD (Figure 3A). Evidence from GWAS, TWAS and other studies show that some genes appear to be risk factors for multiple disorders including *KLC1* for SZ, MDD and ASD (Figure 3B).

**Figure 3.**
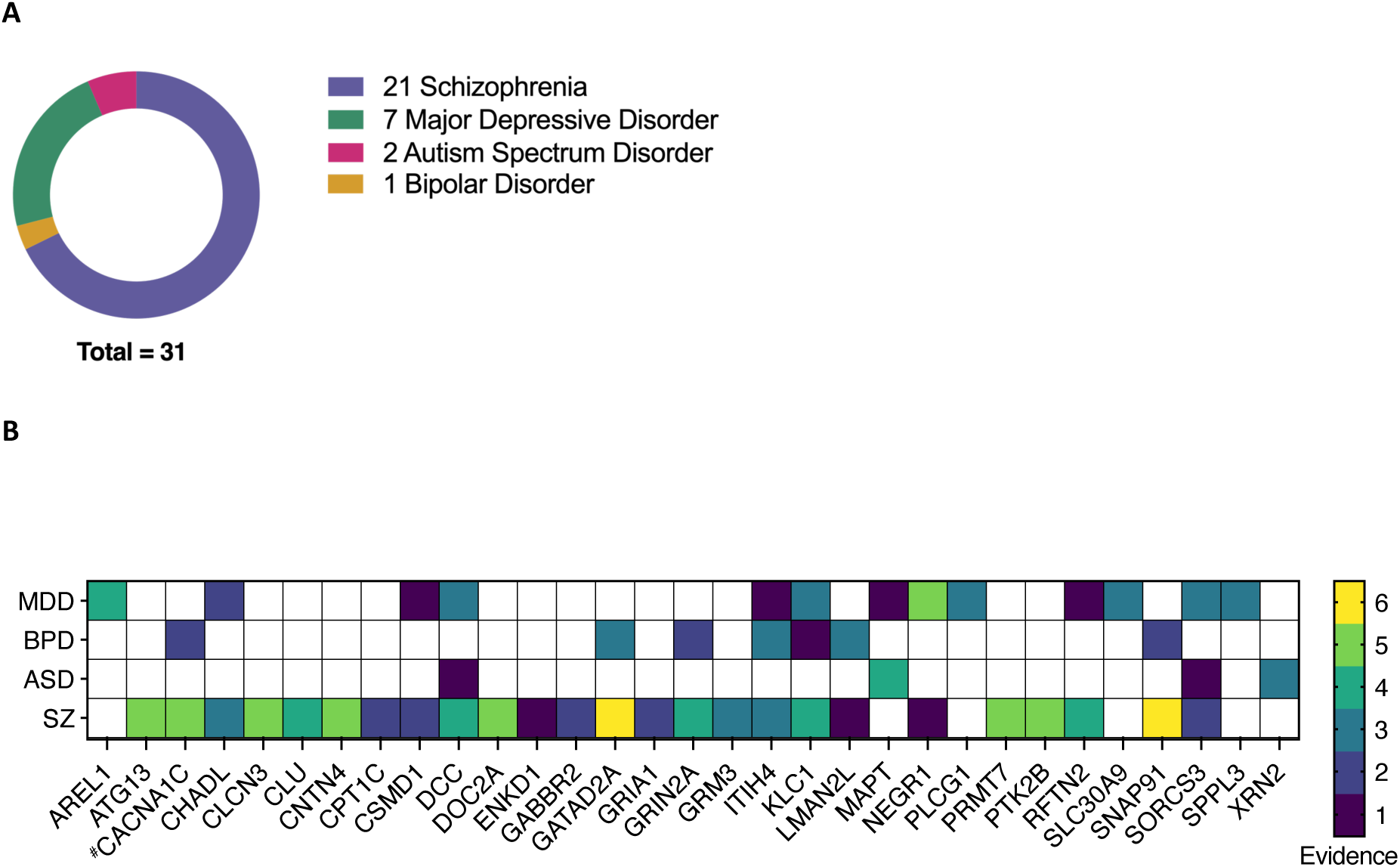
Selection of high-confidence MHD risk genes for amplicon sequencing. **A.** Risk genes included in this study classified by the disorder for which they have the highest evidence of association. **B.** Sequenced genes and their evidence levels for each MHD. The evidence count was calculated as the sum of independent analysis types for example, GWAS, MAGMA, TWAS, SMR, DNA methylation, fine mapping, protein-protein interaction and targeted validation studies, that supported gene involvement in risk for a particular disorder. *^#^*Indicates re-sequencing of a gene from a previous study (Clark *et al.* 2019).

The full RNA isoform profile for each gene was sequenced using nanopore long-read amplicon sequencing. Mapping accuracy ranged from 93.7% (*CLCN3*) to 97.5% (*SORCS3*) (Supplementary Figure 6A, B). Each novel isoform and its predicted impact on known protein domains, open reading frame (ORF) and associated instability index was recorded (Additional File 1) and visualised using IsoVis (Additional File 2) (Wan et al., 2024, *under review*). With no TPM filter set in IsoLamp we identified 872 known and novel isoforms across all 31 neuropsychiatric disorder risk genes. To filter this list for more highly expressed novel isoforms we applied a TPM filter which resulted in 441 known and novel isoforms across all genes (Figure 4A). Of these, SQANTI [42] classified 78 as known (full splice match (FSM)), 256 as novel but using known splice sites or junctions (novel in catalogue (NIC)), and 107 as containing at least one novel splice site (novel not in catalogue (NNC)) (Figure 4A).

**Figure 4.**
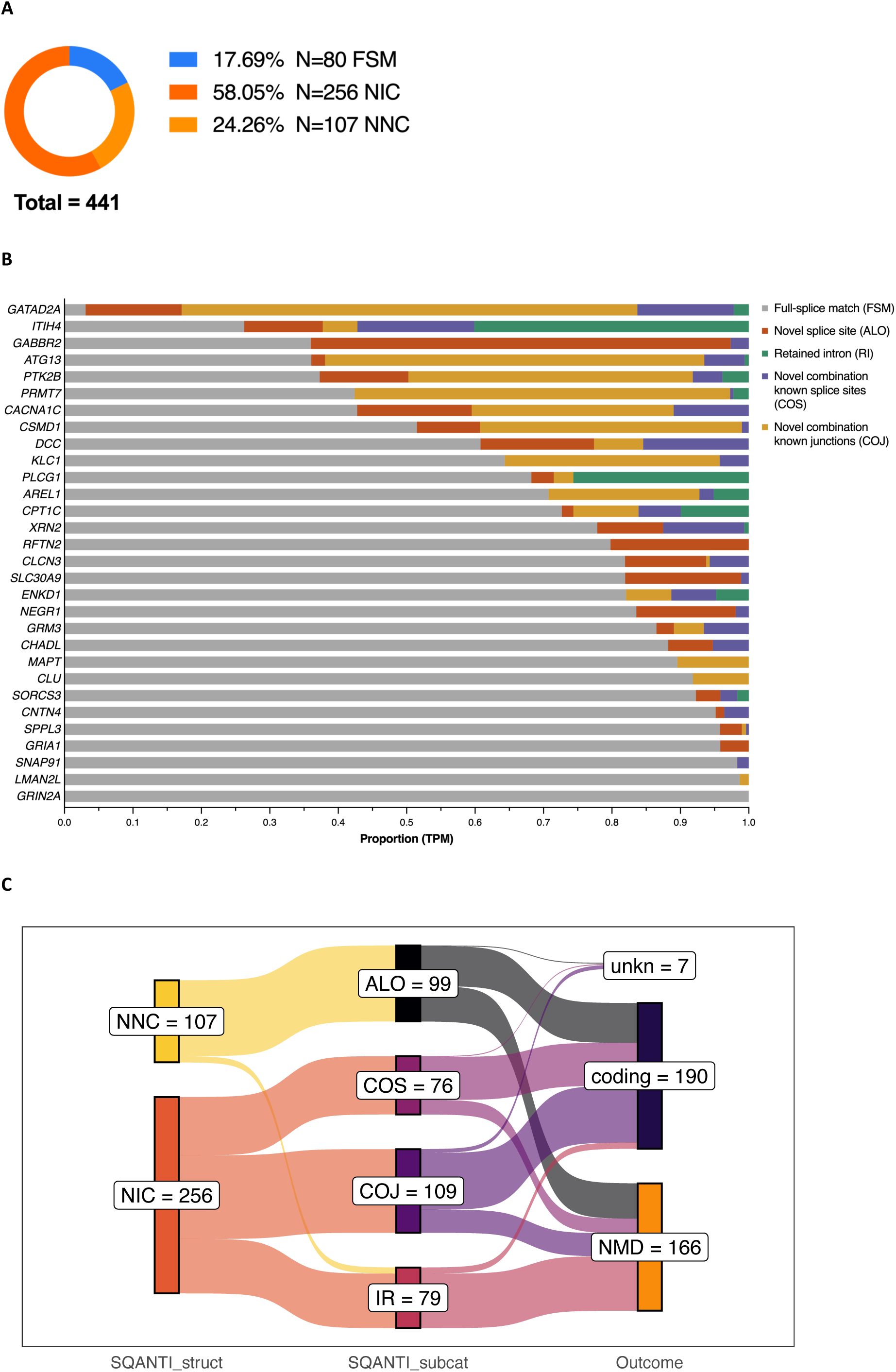
**A.** The total number of known and novel isoforms identified across all 31 risk genes. SQANTI structural categories are known/full splice match (FSM), novel in catalogue (NIC) and novel not in catalogue (NNC). **B.** Proportion of reads (reported as transcript per million, TPM) for each gene as classified by the SQANTI sub-category. **C.** Count of predicted outcomes for novel isoform subcategories. Expasy [55] was used to examine the open reading frame (ORF) of novel isoforms (SQANTI structural category: novel not in catalogue (NNC) or novel in catalogue (NIC)) using the canonical start and stop as a reference. Predictions were categorised as coding if the ORF was retained, nonsense mediated decay (NMD) if a premature termination codon was present and not within 50 nt of the final exon junction or unknown (unkn) if there was not enough information. Novel isoform SQANTI subcategories (subcat) are, at least one novel splice site (ALO), intron retention (IR) and combination of known junctions (COJ) or splice sites (COS).

We next asked what proportion of reads for each gene were assigned to novel isoforms (Figure 4B). This ranged widely from approximately 96.9% for *GATAD2A* to 0% for *GRIN2A*, which was the only gene for which no novel RNA isoforms were detected. Approximately one quarter (7/31) of genes investigated had most of their gene expression assigned to novel isoforms, demonstrating how isoforms and their expression profiles for many genes are still poorly understood. As our amplicon-sequencing does not encompass all variations in transcriptional initiation and termination sites, these results can be seen as a lower bound for the number of novel isoforms. Linear regression of gene isoform counts (Supplementary Figure 7) and novel isoform proportion did not reveal a significant relationship with amplicon length or canonical exon count, indicating that detection of novel isoforms is largely gene dependent (Supplementary Figure 8A-D). To determine what was different about the splicing pattern of each novel isoform we further sub-classified them using SQANTI, based on the use of a combination of known exon junctions (COJ) or splice sites (COS), retained intron (RI) or containing at least one novel splice site (ALO) (donor, acceptor or pair) [42]. Overall, the most reads were assigned to “novel combination of known junctions”, where all individual exon combination were known but the entire chain of exons was novel. The type and proportion of novel isoforms from each category was highly gene specific, demonstrating a wide variety of novel RNA types missing from current gene annotations.

The impact of each novel isoform on the encoded ORF was examined using Expasy [43] and recorded as retaining the canonical or other known reading frame (coding), likely-NMD or unknown. Novel isoforms were classified as coding for 54.2%, 67.3% and 75.4% for ALO, COJ and COS subcategories respectively. We identified 49 novel isoforms that contained retained introns, 39 (83%) of which were predicted to lead to NMD (Figure 4C). Our results were also useful for several genes in identifying the probable isoforms represented by GENCODE transcript fragments. For the SZ risk gene clusterin (*CLU*), the novel Tx1 (COJ) extended ENST00000520796 to the canonical stop codon and suggested this isoform is moderately abundant (8.2% of TPM) across all brain tissues. The ASD risk gene microtubule associated protein tau (*MAPT*) novel Tx5 extended ENST00000703977 and further demonstrated that inclusion of canonical exon 7 (chr17:45,989,878-45,990,075) does not always exhibit coordinated splicing with canonical exon 5. This isoform had moderate expression comprising 3.2% of *MAPT* TPM.

Isoforms that contained ‘at least one novel splice site’ (ALO) generally contained a novel deletion within a known exon or had novel donors and/or acceptors. All novel junctions in ALO isoforms were canonical GT-AG, GC-AG or AT-AC junction pairs, though often only the splice donor (GT) or acceptor (AG) was novel, for example a novel splice acceptor (+98 nt) for *CPT1C* exon 17 (Supplementary Figure 9A, B). We found that ∼48% of ALO isoforms contained either a single novel splice donor or acceptor. Novel GC-AG pairing was detected in two SZ risk genes, within the 5’UTR of *GABBR2* and the donor site of a validated novel exon in *RFTN2*. These results show a clear advantage of using long-read sequencing to contextualise novel splice sites which aids in predicting the outcome on the isoform and ORF.

### Detection of highly expressed novel isoforms

A key question regarding novel isoforms is whether they are expressed at high enough level to impact the biological function of a gene. This is a complex question, because a novel isoform could be low at the tissue level but highly abundant in a specific cell type, or multiple expressed novel isoforms can be significant cumulatively, especially if they all encode the same change to a protein. Therefore, we focused on genes with significant individual or cumulative expression of novel isoforms (analysis on all gene isoforms is available in Additional File 2).

We identified 22 novel isoforms for the schizophrenia risk gene autophagy-related protein 13 (*ATG13*). Novel isoforms represented 64% of gene expression, compared to 36% for full-splice matches. The most abundant class of novel isoforms (15/22) were COJ, which made up 55.4% of gene expression (Figure 4B). *ATG13* had two alternative splicing hotspots, firstly with the 5’UTR and secondly around a predicted disordered region involving exons 12 and 13 in the canonical isoform.

Across all brain regions the most highly expressed isoform was the novel COJ transcript 26 (Tx26), which represented 23% of TPM, surpassing the canonical transcript ENST00000683050 (12.8%). Tx26 differs from the canonical transcript by skipping of exon 12 (Figure 5A, B). It contains the same CDS as ENST00000359513 but includes an additional exon in the 5’UTR (exon 3). Novel COJ transcripts 6 and 8 also had high TPMs and together accounted for 16.6% of expression. These isoforms were novel due to a combination of 5’UTR exons not previously seen within full-length GENCODE annotations.

**Figure 5.**
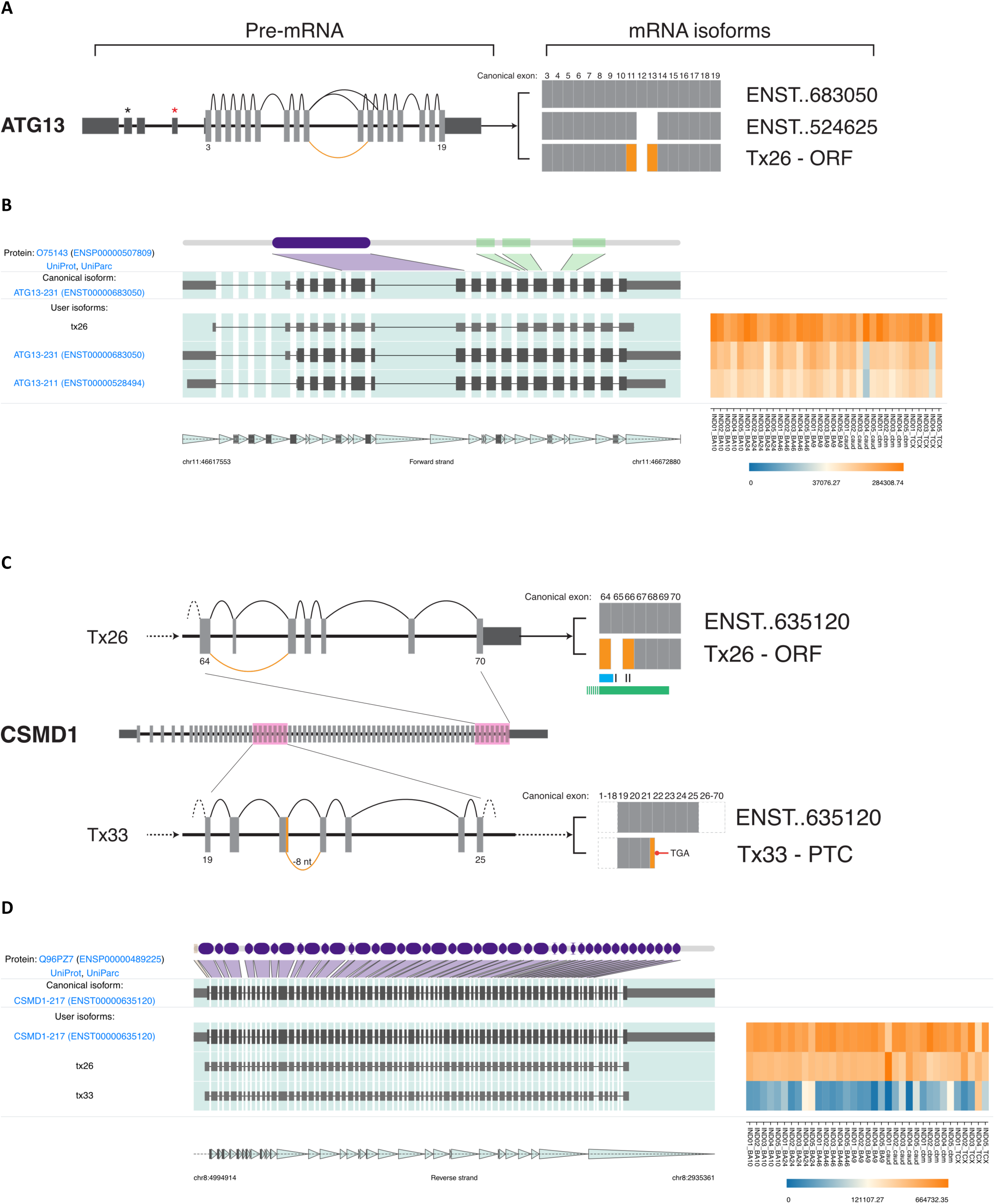
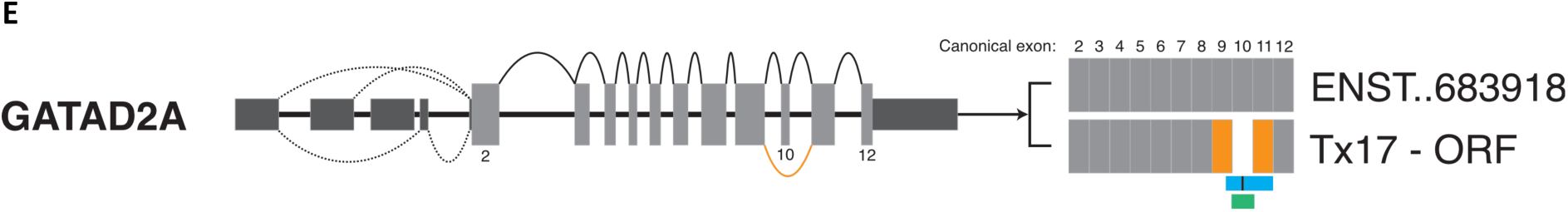
Highly abundant novel isoforms and the predicted mRNA outcome. **(A,C,E)** mRNA splice graphs. Dark and light grey boxes indicate 5’ and 3’ UTR and coding exons respectively. Numbers indicate the coding exon of interest. Orange arcs (pre-mRNA) and boxes (mRNA) indicate novel splicing events. mRNA isoforms depict known isoforms (ENST) against novel (Tx) isoforms, “..” indicates abbreviated zeroes. **(B,D)** IsoVis visualisation of isoform structures (centre stack) and expression levels (heatmap). Canonical isoform shown at top of stack including exonic mapping of protein domains (purple) and disordered regions (green) **A.** Splice graph of *ATG13* highlights the open reading frame (ORF) preserving skipping event of canonical exon 12. **B.** High expression of *ATG13* novel transcript 26 (Tx26). **C.** Splice graphs highlighting novel changes in *CSMD1* novel transcript 26 (Tx26) and 33 (Tx33) within highlighted pink regions. The ORF retaining skipping event of canonical exon 65 may disrupt a known glycosylation site (black bar), a sushi domain extending from exon 64 (blue) and part of an extracellular domain (green). Tx33 contains a novel splice donor (-8 nt) within exon 21 leading to a premature termination codon (PTC) in exon 22. Dashed lines indicate continuation of the transcript to 5’ or 3’ coding exons. **D.** Relatively high expression of *CSMD1* novel transcripts 26 and 33. **E.** *GATAD2A* novel transcript Tx17 contained a novel, ORF retaining, skipping event of canonical exon 10 which contains a phosphorylation site (black bar), part of a polar biased region (blue) and overlaps a CpG island (<300 bp, green). Dashed lines indicate alternative splicing of 5’UTR exons.

The schizophrenia risk gene CUB and sushi multiple domains 1 (*CSMD1*) was the longest CDS we amplified at approximately 10,838 nt encompassing 70 coding exons. In total, 8/9 detected isoforms were classified as novel (Additional File 2I). Following the canonical isoform (ENST00000635120, 51.5% of TPM), novel transcripts 26 (COJ) and 33 (ALO) accounted for 38.3% and 7.1% of isoform TPMs, respectively (Figure 5C, D). Novel Tx26 skipped known exon 65 which encodes a sushi 28 extracellular domain and glycosylation site. The ORF of Tx26 retained the reading frame encoding a 3549 amino acid (aa) protein. Novel Tx33 contained a novel splice donor (GT, -8 nt) in canonical exon 21 and was predicted to encode a PTC in canonical exon 22. The full Tx33 mRNA also skipped canonical exon 65. *CSMD1* also provides a useful example of the benefit of long read for profiling isoforms. GTEx isoform expression data (https://www.gtexportal.org/home/gene/CSMD1) for *CSMD1* in brain is almost exclusively assigned to isoforms with downstream transcriptional initiation sites (including the two-exon fragment ENST00000521646), despite splice junction level expression largely supporting expression from the canonical start site. This emphasises the difficulty of assembling and quantification expression of full-length isoforms from long, complex genes, which can be achieved using long isoform spanning reads.

The chromatin remodelling subunit and shared SZ and BPD risk gene GATA zinc finger domain containing 2A (*GATAD2A*) had one of the highest proportions of reads (96.9%) assigned to novel isoforms (Additional File 2N). Most novel isoforms were predicted to be coding COJs (10/24) and these also accounted for the majority of novel expression (66.6%). Novel Tx17 had the highest expression level (22.7%) of any *GATAD2A* isoform and skipped canonical exon 10 which overlaps a CpG island (212 nt, 21.7% CpG) and contains a disordered, polar residue biased region and a phosphorylation site (Figure 5E). Two additional novel isoforms (Txs 8 and 12), together accounting for 19.9% of expression, incorporated a known 89 nt 5’UTR exon (ENST00000494516) into full-length isoforms for the first time, clarifying the isoforms expressed from this gene. It is important to note that there are two known start sites for this gene supported by high levels of CAGE reads and human mRNA. The forward primer used in this study was located within the 5’UTR of ENST00000360315 and as such expression levels of alternatively spliced isoforms from ENST00000683918 are not included in this analysis (Supplementary Table 3).

Several additional genes had relatively high levels of at least one novel isoform including the SZ risk genes kinesin light chain 1 (*KLC1*), protein tyrosine kinase 2 beta (*PTK2B*) and protein arginine methyltransferase 7 (*PRMT7*). *KLC1* novel transcript 1, 31.4% of gene TPM, was predicted to retain the canonical ORF and included a known (ENST00000380038) splice acceptor (+75 nt) with an alternative 3’ end (Additional File 2S). *PTK2B* novel transcripts 25 and 11 together accounted for ∼42% of total expression and both had variable splicing, including a novel donor (GT, +141nt), of the 5’UTR exon 5 (ENST00000519650) (Additional File 2Y). *PRMT7* novel transcript 18, 12.3% of TPM, skips canonical exon 4 which may lead to NMD or alternatively, use a supported (ENST00000686053) translation start site in the following exon (Additional File 2X).

The full RNA isoform profile for the SZ risk gene calcium voltage-gated channel subunit alpha1 C (*CACNA1C*) has previously been reported and was repeated in this study [35]. In total, we identified 5 annotated and 22 novel isoforms. The most highly expressed novel isoform identified in our previous study, ‘novel 2199’, now known as ENST00000682835.1 was also identified in our samples and exhibited similar cerebellar specific expression. Importantly, 10 novel isoforms replicated one of two alternative splicing events in a hotspot identified previously in canonical exon 7 [35]. This hotspot contains the canonical splice site and two alternative 3’SS acceptors over only 12 nucleotides (chr12: 2,493,190-2,493,201) (Supplementary Figure 10).

### Novel isoforms alter predicted protein structures

Novel isoforms have the potential to affect either post-transcriptional regulation and/or protein sequence, structure and function. We next investigated a selection of isoforms that would be predicted to lead to protein changes to understand their possible impact.

Several novel isoforms (including 5 of the top 20 by expression, Additional File 2R) predicted a novel exon 22 skipping event in the SZ and MDD risk gene *ITIH4*. Targeted mass spectrophotometry confirmed a novel junction between exons 21 and 23 (ETLFSVMPG//PVLPGGALGISSSIR) created due to skipping of exon 22 (Figure 6A, Supplementary Figure 11). This event was predicted to encode a PTC <50 nt from the final exon junction, indicating it may not be directed to NMD. Protein structure prediction of the canonical (ENST00000266041.9) and a representative novel isoform (Tx71) indicated a loss of 106 aa (∼44%) of the 35 kDa heavy chain domain but retention of three O-glycosylation sites (Thr:719, 720, 722) (Figure 6B-D). Novel transcript 71 accounted for ∼3.7% of *ITIH4* TPM and this skipping event was found in an additional 24/68 (∼35%) novel isoforms which together accounted for 23.4% of TPM. Tx71 also skipped canonical exons 15 and 16, which contain a protease susceptibility region (residues 633 - 713) and a MASP-1 cleavage site (645 - 646: RR) [44]. Cleavage at this site and subsequent formation of an ITIH4-MASP complex can inhibit complement activation via the lectin pathway [44]. However, skipping of canonical exon 22 was not mutually inclusive with skipping of exons 15/16 as other novel isoforms with exon 22 skipping retained exons 15/16. The absence of much of the 35 kDa heavy chain domain is likely to impact on ITIH4 protein function and further studies will be required to examine if it plays a role in neuronal phenotypes.

**Figure 6.**
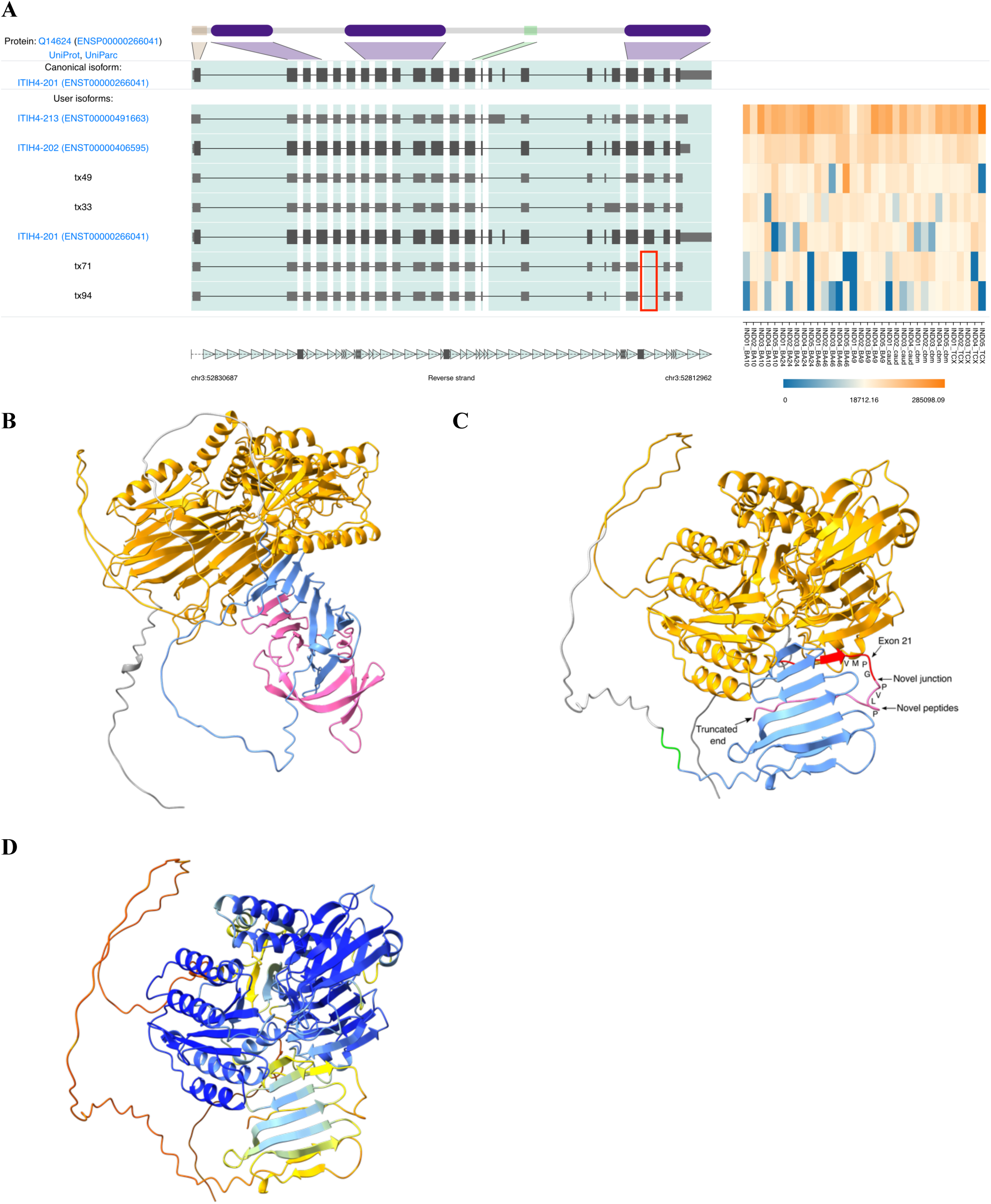
*ITIH4* canonical and novel isoform protein structure predictions. **A.** IsoVis stack of the top seven *ITIH4* isoforms sorted by expression. Several novel isoforms contained the novel exon 22 skipping event (red box) including Txs71 and 94. **B.** Canonical isoform (ENST00000266041, UniProt:Q14624) structure prediction indicating 70 kDa (orange) and 30 kDA (blue) chains **C.** Novel isoform (Tx71) structure prediction indicating 70 kDa chain (orange), truncated 30 kDA chain (blue), O-glycosylation sites (green), novel splice junction peptide detected using mass spectrophotometry (red) and novel peptides (pink). Black arrow indicates termination <50 nt from the final exon junction complex. **D.** AlphaFold per-residue confidence scores (pLDDT) (0-100) for *ITIH4* novel transcript 71: very high (>90, blue), confident (90-70, light-blue), low (70>50, yellow) and very low (<50, orange).

Ten novel isoforms were identified for the SZ risk gene glutamate metabotropic receptor 3 (*GRM3*). Three novel isoforms (Txs 6, 7, 9) skip exon 2 which contains the canonical translation start site and instead could use an alternative, frame retaining, translation initiation site in exon 1, extending the truncated reference isoform ENST00000454217.1 which is also supported by human amygdala mRNA (AK294178) (Additional File 2Q). Translation of these isoforms would likely cause significant disruption to the resultant protein with removal of the signal peptide, transmembrane domain and disulphide bonds. Cumulatively these novel isoforms accounted for a relatively low 8.8% of expression when compared to the canonical isoform (86.5%).

Both novel isoforms and exons were identified for the shared MDD and ASD risk gene neuronal growth regulator 1 (*NEGR1*). Most reads (83.5%) were assigned to the canonical *NEGR1* isoform (ENST00000357731, 354 aa). Three novel isoforms were identified, two of which (Txs 1 and 2) contained novel exons (Figure 7A, B). These transcripts accounted for 9.4% (Tx2) and 5.1% (Tx1) of TPM. Both novel exons were located between cassette exons 6 - 7 and were validated using Sanger sequencing. The novel exon within Tx1 was 42 nt (14 aa) in length, had high 100 vertebrate conservation (UCSC) and was predicted to be frame retaining (Supplementary Figure 12A). Protein structure prediction of the 368 aa Tx1 using AlphaFold [45] showed a 14 aa extension near the C-terminal prior between the GPI anchor (G:324 aa) and the three immunoglobulin-like domains (Supplementary Figure 12B). In contrast, the 58 nt novel exon within Tx2 encoded a PTC (TAG) only 35 nt distant to the final exon junction complex, suggesting it might not trigger NMD. Truncation of the protein at this position (313 + 7 novel aa) would remove the GPI anchor potentially creating a near complete protein (320 aa) that is unable to attach to the cell membrane (Supplementary Figure 12C).

**Figure 7.**
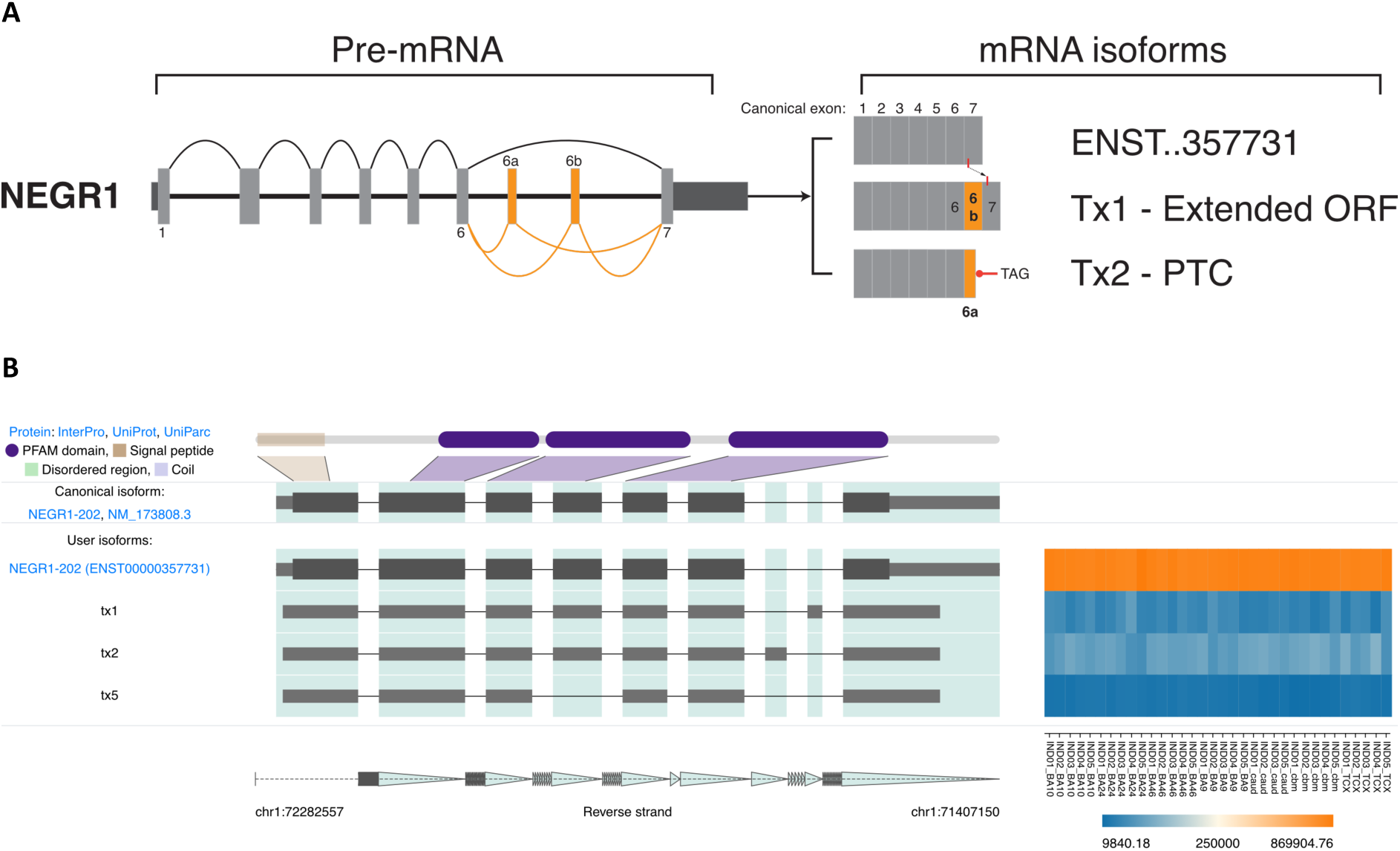
*NEGR1* splice isoforms in human brain. **A.** *NEGR1* mRNA splice graph highlighting validated novel exons 6a and 6b. Dark and light grey boxes indicate 5’ and 3’ UTR and coding exons respectively. Numbers indicate the coding exon of interest. Orange arcs (pre-mRNA) and boxes (mRNA) indicate novel splicing events/exons. mRNA isoforms depict known isoforms (ENST) against novel (Tx) isoforms, “..” indicates abbreviated zeroes. In the open reading frame (ORF) retaining Tx1, a GPI anchor (red line) is shown to shift 3’ in the final exon when compared to ENST00000357731. Tx2 encodes a premature termination codon (PTC) within the novel exon. **B.** IsoVis visualisation of *NEGR1* isoform structures (centre stack) and expression levels grouped by brain region (heatmap). Canonical isoform shown at top of stack including exonic mapping of a 5’ signal peptide (brown) and three immunoglobulin (Ig)-like domains (purple). Canonical 3’UTR has been trimmed.

### Brain region specific expression of novel isoforms

Many isoforms have brain region enriched or specific expression [46, 47]. Our amplicon sequencing approach identifies the presence and relative expression proportion of different isoforms. We next asked if any novel risk genes isoforms showed expression differences between brain regions. Overall, cerebellum exhibited most differences in isoform expression, consistent with previous whole transcriptome results [12].

Depression risk gene DCC netrin 1 receptor (*DCC*) novel isoform Tx9 had significantly higher TPM in cerebellum (Figure 8A, Additional File 2J). TPMs of Tx9 in CBM were approximately 10x higher than the average for cortical regions and 3x higher than in caudate. This isoform, classified as a COJ and predicted to encode a 1425 aa protein, accounted for ∼5% of total *DCC* expression. Tx9 uses an alternative 3’SS (-60 nt) in cassette exon 17 and the skipped nucleotides cover an extracellular region and fibronectin type-III domain (UniProt). The SZ risk gene double C2 domain alpha (*DOC2A*) had two novel isoforms with significant variation in brain specific expression including Tx8 in cerebellum and Tx53 in caudate (Figure 8B, C, Additional File 2K). Novel Tx8 used a novel splice donor in canonical 5’UTR exon 1 (GT, +158 nt) and was predicted to encode a 400 aa protein unchanged from the canonical transcript. Tx53 was the only novel transcript that showed moderate but specific expression in caudate samples or any tissue other than cerebellum. Tx53 extends the known isoform ENST00000574405 to the canonical stop and is predicted to encode a 400 aa protein. Overall, 28 novel isoforms in 11 risk genes were found to have variable expression amongst brain tissues supporting a role for these isoforms within specific brain regions or potentially in a subset of cells.

**Figure 8.**
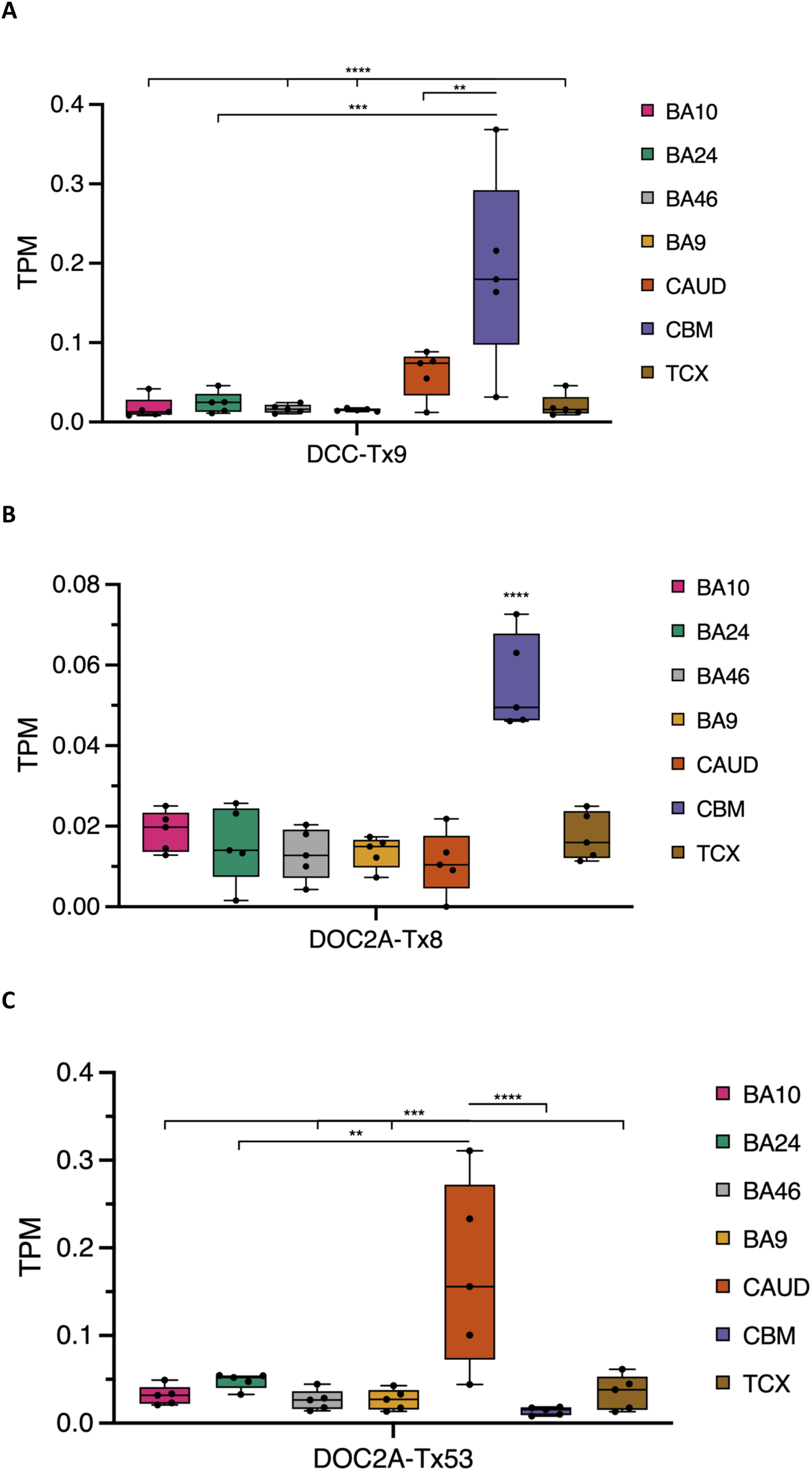
Brain region specific expression of novel isoforms. **A.** *DCC* novel transcript 9 (Tx9) uses a known (ENST00000581580) alternative 3’SS (-60 nt) in cassette exon 17 and had significantly higher TPM in CBM. ANOVA: F=9.098, DF=28. **B-C.** *DOC2A* novel transcripts. Tx8 used a novel splice donor in canonical 5’UTR exon 1 (GT, +158 nt). ANOVA: F=20.73, DF=28. Tx53 extended the known reference isoform ENST00000574405 to the canonical translational stop and had significantly higher TPM in caudate. ANOVA: F=8.261, DF=28. Brodmann’s Area (BA), caudate (CAUD), cerebellum (CBM) and temporal cortex (TCX). Ordinary one-way ANOVA Tukey’s multiple comparison adjusted P value: **= P ≤ 0.01, ***=P ≤ 0.001, ****=P ≤ 0.0001.

### Sequencing and validation of novel exons

Our amplicon sequencing approach detected a total of 28 novel exons in 13 MHD risk genes. Using RT-PCR followed by Sanger sequencing we validated a set of 21/21 targeted novel exons (Table 1). The SZ risk gene chloride voltage-gated channel 3 (*CLCN3*) contained four novel exons within six novel isoforms, and an example of PCR validation is shown in Supplementary Figure 13A. Validated novel exon mean length was 99 nt, ranging from 41 nt (*CLCN3*) to 231 nt (*GRM3*). 16 (76%) of validated novel exons were classified as ‘poison exons’ as they encoded a PTC (Supplementary Figure 13B), although two of these poison exons, within *NEGR1* and *XRN2*, were <50 nt from the final exon junction and therefore may not undergo NMD. The novel exon contained within Tx3 for *XRN2* had the second highest isoform expression for the gene, following the canonical transcript (ENST0000037191), with 4.7% of TPM. If translated, this transcript would omit an omega-N-methylarginine modification site (ARG:946) within a disordered region at the C-terminus (Supplementary Figure 14, Additional File 2AE).

**Table 1.** Neuropsychiatric disorder risk gene novel exon validation. *<u>Definitions</u>: chromosome (Chr), nucleotide (nt), open reading frame (ORF), premature termination codon (PTC), untranslated region (UTR)*.

| Gene | Novel exon | Chr | Genomic coordinates |  | Size (nt) | Classification | UniProt/Pfam domain |
| --- | --- | --- | --- | --- | --- | --- | --- |
|  |  |  | Start | End |  |  |  |
| *SORCS3* | 20a | 10 | 105244778 | 105244837 | 60 | ORF | Lumenal |
| *GRIA1* | 2a | 5 | 153509750 | 153509806 | 57 | PTC | Extracellular |
| *XRN2* | 16a | 20 | 21345834 | 21345887 | 54 | PTC | None |
| *XRN2* | 29a | 20 | 21387308 | 21387371 | 64 | PTC (<50nt) | Disordered |
| *SLC30A9* | 6a | 4 | 42028140 | 42028260 | 121 | PTC | Helix |
| *SLC30A9* | 9a | 4 | 42059964 | 42060036 | 73 | PTC | Cation efflux family |
| *GRM3* | 2a | 7 | 86776784 | 86777014 | 231 | PTC | None |
| *GRM3* | 3a | 7 | 86832296 | 86832411 | 116 | PTC | None |
| *NEGR1* | 6a | 1 | 71532866 | 71532907 | 42 | ORF | None |
| *NEGR1* | 6b | 1 | 71587343 | 71587400 | 58 | PTC (<50nt) | Transmembrane |
| *RFTN2* | 1a | 2 | 197654257 | 197654391 | 135 | PTC | None |
| *RFTN2* | 1b | 2 | 197671542 | 197671664 | 123 | PTC | None |
| *RFTN2* | 6a | 2 | 197616952 | 197617073 | 122 | PTC | None |
| *CNTN4* | 1a | 3 | 2656485 | 2656587 | 103 | PTC | None |
| *PTK2B* | 3a | 8 | 27318676 | 27318793 | 118 | UTR | None |
| *PTK2B* | 3a (short) | 8 | 27318696 | 27318793 | 98 | UTR | None |
| *PTK2B* | 3b | 8 | 27319918 | 27319985 | 68 | UTR | None |
| *CLCN3* | 1a | 4 | 169630165 | 169630335 | 171 | UTR | None |
| *CLCN3* | 2a | 4 | 169638597 | 169638742 | 146 | PTC | Cytoplasmic |
| *CLCN3* | 2b | 4 | 169640168 | 169640208 | 41 | PTC | Cytoplasmic |
| *CLCN3* | 2c | 4 | 169663527 | 169663617 | 91 | PTC | Cytoplasmic |

Three novel exons were in untranslated regions and two were predicted to retain the ORF, including the 42 nt exon in *NEGR1* mentioned previously and a 60 nt exon within *SORCS3*. *SORCS3* is a member of the VPS10 transmembrane protein family and assists with neuronal protein trafficking and sorting and a lack of *SORCS3* in the hippocampus in mice has been associated with impaired learning and fear memory in mice [48–50]. The novel exon in Tx1, encoding 20 aa (AMCGRAQWFTPVILALWETE), falls within the *SORCS3* lumenal region (position: LYS:956/PRO:957) and did not appear to disrupt the transmembrane or cytoplasmic domains.

Comparison of protein prediction models of the canonical (ENST00000369701.8) and novel isoform (Tx1) showed the addition of an unstructured loop with a partial alpha helix within the second polycystic kidney disease (PKD2) domain (Figure 8A-D), though the prediction confidence was low, so the structural impact on the PKD2 domain remains uncertain [51].

## Discussion

In this study we used long-read sequencing to profile 31 neuropsychiatric disorder risk genes identifying 363 novel RNA isoforms. We also present a new bioinformatic tool, IsoLamp, that can accurately identify and quantify novel RNA isoforms from long-read amplicon data. The recent proliferation of GWAS studies examining increasingly large population-wide data has identified hundreds of genomic variants associated with risk of developing a mental health disorder [23, 52]. Evidence suggests that some risk variants, specifically those that are non-coding, play a role in pre-mRNA splicing and our current understanding of the transcriptomic profile for these risk genes is limited [35, 53]. A key finding of our research is both the high number of novel expressed RNA isoforms and, for some risk genes, the high expression of novel isoforms both individually and collectively. This finding reflects both the known complexity of alternative splicing in the human brain [54] and the current incompleteness of the reference transcriptome. As a result of the relatively deep sequencing afforded by this long-read approach, we have shown that there is a much higher level of RNA isoform diversity for these genes than reported in the current reference annotations. These findings provide new insight into the repertoire of RNA isoforms expressed in brain that could be important for understanding the risk and onset of neuropsychiatric disorders.

### RNA isoform discovery, classification and visualisation

We generated a set of high-confidence RNA isoforms from nanopore long-read data using IsoLamp. IsoLamp optimises and streamlines transcript identification, quantification and annotation from long-read amplicon data and outperformed other methods. IsoLamp improves upon our previously published pipeline TAQLoRe [35] by expanding analysis capabilities to any gene and identifying all isoforms within a single pipeline. Our overall approach also overcomes the significant challenge of re-assembling and classifying RNA isoforms using short reads [55–57]. The primary outputs from IsoLamp, filtered transcripts (GTF) and transcript expression (TPM) is designed to be compatible with multiple downstream tools, including our visualisation tool IsoVis (https://isomix.org/isovis) (Wan et al., 2024, *under review*).

Taken together, our benchmarking results highlight that IsoLamp’s optimised isoform discovery parameters, coupled with its specific filters (expression, overlapping primers, and full-length reads), yield significant improvements in both precision and recall compared to Bambu, FLAIR, FLAMES and StringTie2. The TPM filter applied to the data presented in this study is conservative and for long and complex genes may need to be tested to yield a balance of novel isoform detection and acceptable expression levels. IsoLamp also output consistent expression quantification that was robust to the quality of the annotations provided.

### Novel RNA isoforms in neuropsychiatric disorders

The results presented in this study confirm our current limited understanding of RNA isoform profiles in the human brain and demonstrates how long-read sequencing is a powerful tool to address this issue [34, 35, 58].

We identified several highly abundant novel isoforms, including one, *ATG13* (Tx26), that was the most abundant gene isoform. *ATG13* forms part of a protein complex, including ULK1 and FIP200, that is critical for autophagy [59]. In our samples, transcript 26 had the highest TPM and contained a known skipping event of canonical (ENST000006683050) exon 12 which may be involved with FIP200 binding and subsequent function of ATG13 [60]. Where Tx26 and several other novel isoforms differ from known (e.g. ENST00000359513) isoforms is in the 5’UTR, indicating this region may play a role in translation regulation [61]. Similarly, the SZ risk gene *CSMD1*, had relatively high expression of novel isoforms. Recent evidence suggests enrichment of CSMD1 protein in the brain and activity as an inhibitor of the complement pathway in neurons [62]. Baum *et al*. (2024) show that CSMD1 localises to synapses and that loss of CSMD1 can lead to increased complement deposition potentially disrupting complement associated synaptic pruning. The novel isoforms identified in our samples provide transcriptional pathways through which *CSMD1* may be altered, potentially reducing expression or function of the protein, for example through incorporation of a premature termination codon (Tx33). Evidence from Alzheimer’s disease studies has also linked increased complement pathway activity to cognitive impairment, however further studies, particularly in human models of neuronal development will be needed to link *CSMD1* transcriptional variability to SZ risk and severity [62, 63]. *KLC1* encodes one of the two light chains required for the microtubule motor protein kinesin, mutations in which have previously been found in SZ patients and which resulted in a SZ-like phenotype in mice [64]. Novel *KLC1* Tx1 (∼31% TPM) was predicted to alter the C-terminal end of the protein, an area known to undergo alternative splicing to expand cargo binding diversity [65]. The combination of an altered C-terminal tetratricopeptide repeat (TPR), 3’UTR and relatively high expression suggests novel Tx1 plays a role in brain, though the functional impact on the protein requires further validation.

Several novel isoforms and exons were identified for the ion homeostasis and channel genes *CLCN3*, *CACNA1C* and *SLC30A9* which have shared risk for SZ, BPD and ASD [21, 23, 52]. Voltage-gated ion channels are widely distributed in the brain and regulate neuronal firing. Mutations to these genes have been associated with disease and the emerging role of these channels in neuropsychiatric disorders has been previously reviewed [66]. *CLCN3* belongs to the CLC family of anion channels and transporters and has an established role in human neurodevelopment [67, 68]. We identified and validated four novel exons in *CLCN3*, three of which were predicted to encode a PTC which could lead to NMD. The fourth was located within the 5’UTR, an area known to impact translation regulation in humans potentially through structural interference with the ribosome [61]. Splice variants of *CLCNC3* have been shown to impact intracellular localisation and our results identified additional splice variants, in particular a novel RNA isoform (Tx9) which is similar to ENST0000613795, but includes a 76 bp exon 12 [67]. Twenty-two novel isoforms were identified for calcium channel gene *CACNA1C* and supporting previous findings [35] the top ten novel isoforms, by TPM, were classified as frame retaining, supporting their potential to generate functional proteins.

*SLC30A9* (first known as *HUEL*) encodes the zinc transporter protein 9 (ZnT9), which is involved in zinc transport and homeostasis in the endoplasmic reticulum [69]. Whilst the function of the protein is not fully understood, a 3 nt familial deletion (c.1047_1049delGCA) in the highly conserved cation efflux domain (CED) has been recorded to result in changes to protein structure, intracellular zinc levels and intellectual disability [70, 71]. Critically, we identified and validated a novel exon (Tx3-9a:73 nt) within this CED providing evidence that this region may be alternatively spliced more commonly than previously understood, potentially impacting protein function [69].

Finally, using mass spectrophotometry we confirmed a novel skipping event in the ITIH4 protein, indicating the utility of combining long-read sequencing with proteomics. ITIH4 is an acute-phase protein the serum levels of which have been associated with MDD and is thought to be involved in neuro-inflammation [72, 73]. Despite several studies outlining an association for *ITIH4* and risk for SZ, BPD or MDD onset, the causal mechanisms for this gene remain elusive and a further study is required to further explore the impact of the coding change detected in our study.

### Limitations and future directions

Long-read amplicon sequencing, while providing an extremely sensitive isoform quantification method, is limited by the set(s) of primers used to amplify each risk gene. Our method aimed to locate primers within the 5’ and 3’ UTRs in proximity to the canonical translational start and stop codons.

This approach generally amplified the entire coding sequence but does not capture full-length isoforms or variation in the UTRs and additional primers must be made to capture alternative unique start and termination sites. When using this method users must interpret the reported novel isoform proportions in the context of the known isoforms targeted e.g. if the canonical isoform is not a target of the primer pair, novel isoform expression may appear inflated.

The results of our study are limited by the sample size of available control post-mortem brain tissue. The nanopore long-read data for each risk gene was generated from five elderly, male control individuals, with a single female sample removed from further analyses due to quality constraints.

The small number of available individuals means this dataset was not powered to investigate genotype impacts on isoform expression, though this will be an important area of investigation to determine which risk genotypes act through changes in isoform structure and/or expression.

Sample and RNA quality, as measured by RINe, is critical to high-quality sequencing and this is especially true for long reads [35, 74]. Supporting previous findings in mRNA, our data suggest that pH values <6.3 impacted the quality of post-mortem human brain RNA, which is especially critical for robust amplification of longer (>5 Kb) CDS [75]. In general, PCR cycling was kept as low as possible to avoid PCR bias towards shorter isoforms and other artifacts. However, we noted that lower RINs, as recorded for Ind04, appeared to impact amplicons of longer CDS. To help overcome such issues, future long-read amplicon sequencing could incorporate unique molecular identifiers to tag molecules prior to PCR to ensure an accurate representation of the original RNA isoforms [76].

The risk genes profiled in this study were selected based on multiple levels of evidence for their involvement in risk, not only from GWAS but from meta-analyses and further independent studies [22, 25]. Whilst this approach was expected to produce a set of genes with high-confidence of their involvement in disorder risk, it is not exhaustive and it will be important to ensure risk gene lists are updated as more evidence from GWAS and other studies becomes available [25, 53]. Genes that are thought to confer resistance to the development of neuropsychiatric disease are also beginning to emerge and the addition investigation of their expression and isoform profiles may also provide valuable insight into disease risk and progression [77]. In future, combining whole or amplicon transcriptomic data, large-scale proteomic data and machine-learning predictive models like TRIFID can help to identify and prioritise functional proteomic isoforms [78].

## Conclusion

In conclusion, we identified hundreds of unreported RNA isoforms and novel exons, many of which could impact the function of known neuropsychiatric risk genes that also play crucial roles in normal neuronal development and activity. An understanding of the regulatory and functional impacts of these novel isoforms and incorporating long-read Nanopore data into existing repositories will help form an important knowledge base of alternative splicing in the human brain [79, 80]. Some novel isoforms or exons may also be future therapeutic targets through the modulation of splice isoforms using antisense oligonucleotides or CRISPR technology.

## Methods

### Sample preparation and QC

Healthy control post-mortem human brain samples were obtained from six individuals collected through the Victorian Brain Bank (VBB) under HREC approvals #12457 and #28304. Age, sex and additional details including the post-mortem interval (PMI), pH and tissue weight are shown in Supplementary Table 1. Briefly, samples comprised 5 males and 1 female, age range: 51 – 72 yrs, PMI range: 31 – 64 hrs and pH range: 5.7 – 6.7. Due to low RNA integrity number equivalent (RINe) the female control was removed from further analysis. Frozen tissue (weight range: 57 – 135 mg) was cut from seven brain regions including Brodmann areas (BA), BA9 (dorsolateral prefrontal cortex (DLPFC)), BA46 (medial prefrontal cortex (MPFC)), BA10 (fronto-parietal cortex (FPC)), Brodmann Area 24 (dorsal anterior cingulate cortex (dACC)), caudate, cerebellum and temporal cortex. Total RNA was extracted from bulk tissue in eight randomised batches of 3 – 6 samples. First, frozen brain tissue was homogenised on ice, using a manual tissue grinder (Potter-Elvehjem, PTFE), whilst immersed in 1 mL QIAzol Lysis Reagent (QIAGEN). Lysate was then processed using a RNeasy Lipid Tissue Kit (QIAGEN, 74804), according to the manufacturers’ instructions. Isolated RNA quality and quantity was checked using a Qubit 4 Fluorometer (2 μL), TapeStation 4200 (RINe: cut-off = 6) and Nanodrop 2000.

### Database curation and risk gene selection

MHD risk genes were selected for long-read amplicon sequencing using an internal database that aimed to collate evidence from the literature of gene involvement in disease risk from the original GWAS, meta-analyses including MAGMA (and variants including eMAGMA, hMAGMA, nMAGMA), TWAS, SMR and follow-up studies including fine mapping, protein-protein interaction (PPI), epigenetic (DNA methylation) and targeted experimental validation (Supplementary Figure 1). The foundation of this database was a list of significant GWAS SNPs for SZ, BD, MDD and ASD. Association data was downloaded from the NHGRI-EBI GWAS Catalog [81]. MHD GWAS associations were filtered on the ‘Disease/Trait’ column to exclude effects of treatments including pharmaceutical, mixed disorder studies and associations with behavioural traits like smoking or alcohol intake. Associations were excluded if both the ‘reported gene’ column was ‘not reported (NR)’ and the ‘mapped gene’ column was blank. Date data was downloaded, filters applied, and percentage associations retained are detailed in Supplementary Table 4.

Follow-up studies and experiments were then identified in the literature and the reported genes were manually collated and assigned to an ‘evidence’ category (i.e.: MAGMA, SMR, experimental validation etc). Each evidence category had the following information headers including the PubMed ID of the reporting manuscript, the first author and the reported SNP and gene. An R script was used to curate risk genes and appearances in unique studies. Counts of reported risk genes and unique studies, for each evidence category, were then combined with the original GWAS table. Risk genes were then sorted by evidence (high to low), separately for each MHD. A multi-trait evidence list was also made by combining each MHD table together and again sorting by descending evidence. This gave us flexibility to focus on risk genes that appeared to be specific to a single MHD or those with shared risk across disorders.

### Primer design, cDNA synthesis and long-range PCR

Thirty-one (31) MHD risk genes were selected from our database and the full coding sequence (CDS) from the canonical isoform was downloaded from the UCSC Genome Browser [82]. Primers, located in the 5’ and 3’ UTRs, were designed to amplify the CDS using Primer3 Plus [83]. Additional primers were made to amplify alternative start or end sites that were not captured by a single primer pair. Additional UCSC track sources including expressed sequence tags (EST), transcript support level (TSL), APPRIS designation, human mRNA support, cap-analysis of gene expression (CAGE) peaks, CpG islands and H3K4Me3 marks were examined to ensure there was enough evidence that alternative start or end sites were real before a primer was designed [82]. All primers, primer combinations and modified Primer3 Plus settings are listed in Supplementary Table 3. Risk gene primers from Primer3 Plus were aligned to tracks on the UCSC Genome Browser using BLAT for visualisation and tested using the In-Silico PCR [82].

To amplify risk gene CDS, 1 μg of total RNA was used as template for cDNA synthesis using Maxima H Minus Reverse Transcriptase (Thermo Fisher Scientific, EP0752, 200 U/μL) according to the manufacturers’ instructions. Two duplicate cDNA plates were generated simultaneously to reduce variability and provide enough template for multiple risk gene PCRs. Risk genes were amplified using one of following DNA polymerases; LongAmp® Taq 2X Master Mix (NEB, M0287S), Platinum™ SuperFi II PCR Master Mix (Thermo Fisher Scientific, 12368010) or PrimeSTAR GXL (TakaraBio, R050B). Each set of gene primers were individually optimised by adjusting PCR cycling conditions (Supplementary Table 3) until sufficient pure template (∼1 – 10 ng) could be produced for input to barcoding. Short-fragments and primer-dimer were removed prior to barcoding using AMPure XP beads (Beckmann Coulter) at 0.5 – 0.8x ratios. An overview of the experimental protocol is shown in Figure 1A.

### Long-read amplicon sequencing

Barcoding conditions for sample multiplexing (N=35, EXP-PBC096, ONT) and library preparation for long-read sequencing followed the recommended ligation sequencing protocol (Figure 1A) (SQK-LSK109/110, ONT). All barcoding PCR was done using LongAmp® Taq 2X Master Mix with an amplicon specific extension time (approximately 1 min/Kb) and 10 – 15x cycles. AMPure clean-up following adaptor ligation was adjusted from the default ratio of 0.4X depending on the length of the target amplicon. Adaptor ligated libraries were loaded (25 – 35 fmol) onto MinION (FLO-MIN106) flow cells and a minimum of 10,000 reads per sample were targeted before flushing and storing the flow cell. All runs were re-basecalled using the super-accurate (SUP) basecalling model (Guppy v6.0.17, 2022) and minimum qscore = 10.

### Isoform discovery from long-read amplicon sequencing with IsoLamp

We developed a new bioinformatic pipeline, IsoLamp, for the analysis of long-read amplicon data (Figure 1B). First, pass reads were down sampled [84] to a consistent number (8000) per barcode and mapped to the reference genome with minimap2 (v2.24) [37]. Then, low accuracy reads (≥5% error rate) were removed and samples were merged prior to isoform identification. Read accuracy was calculated using CIGAR strings in the BAM files and is defined as: (‘X’+’=’+’I’+’D’-‘NM’)/(‘X’+’=’+’I’+’D’). Next, the merged BAM file of high accuracy reads was used as input for isoform discovery with Bambu (v3.2.4) using the following parameters: novel discovery rate (NDR) = 1 and min.fractionByGene = 0.001 [38]. Next, isoforms identified with Bambu were filtered to remove any with zero expression and to retain only isoforms overlapping the known primer coordinates using bedtools intersect (v2.30) [85]. Finally, the remaining isoforms were used to create an updated transcriptome with GffRead (v0.12.7) [86], and reads from each barcode were quantified with salmon in alignment-based mode (v0.14.1) [87] and isoforms were annotated with GffCompare (v0.12.6) [86]. The pipeline outputs a list of isoform annotations (.gtf), isoform expression as transcripts per million (TPM) and proportion of overall gene expression, as well as a report summarising the results. An optional TPM filter was also applied to further filter isoforms which required a minimum TPM of 5000. If the user specifies a grouping variable for their input samples, a t-test is performed between isoform proportions between groups and a false discovery rate of 0.05 is applied.

We benchmarked the performance of the IsoLamp pipeline using Spike-in RNA Variant (SIRV) Set 1 synthetic RNA controls (Lexogen). SIRV isoforms are present in three mixes (E0, E1, E2) that contain isoforms in varying known concentrations. Primers were designed to amplify from the first to the last exon (as described above) of SIRV5 and SIRV6 genes from cDNA generated in triplicate from each SIRV mix (N=27) (Supplementary Figure 2). PCR amplification conditions for SIRV amplicons are shown in Supplementary Table 5. Samples were barcoded and sequenced as described above, and subsequent basecalling and demultiplexing was performed with Guppy (v6.0.17, SUP, 2022). The IsoLamp pipeline was compared against four other isoform discovery tools: StringTie2 [41], FLAIR [39], FLAMES [40] and Bambu [38] (using both default parameters and the optimised parameters used in IsoLamp). The sensitivity, specificity, accuracy and correlations (expected versus observed counts) of the programs were compared using three SIRV reference annotations: complete (C), incomplete (I) (missing isoforms, to test ability to recover unannotated true positive isoforms) and over (O) (extra isoforms, to test ability to minimise false positive annotated isoforms). Novel isoforms were categorised using SQANTI (v4.2) against the human reference (GENCODE release 41, GRCh38.p13). Finally, a combined dataset of expression values for each known and novel isoform and its associated metadata including brain region, gene, RINe, pH, individual, PMI and age was analysed using principal component analysis (PCA).

### Novel exon validation

Nanopore long-read supported novel exons were validated by RT-PCR. Amplicons were designed from the known 5’ flanking exon into the novel exon and from the novel exon into the known 3’ flanking exon. An amplicon spanning the known 5’ and 3’ flanking exons was used as a positive control. Primers were designed using Primer3 [83] and checked using Primer BLAST [88] and are listed in Supplementary Table 6. In some cases, the primer design space was restricted by the novel exon sequence length and/or nucleotide composition. Novel exons were amplified using *Taq* 2X MasterMix (NEB, M0270L) and cycling conditions can be found in Supplementary Table 6. PCR products were visualised via gel electrophoresis using GelGreenⓇ Nucleic Acid Stain (Biotium, 41005) and GeneRuler 100 bp ladder (TFS, SMN0243). PCR products in the expected size range were cleaned up using AMPure XP Reagent (Beckmann Coulter, A63881) at a 1.8X ratio to remove fragments <100 bp and sent for Sanger sequencing (100 – 200 bp, AGRF).

### Protein isolation and novel sequence detection using targeted mass spectrophotometry (MS)

Post-mortem frontal cortex and cerebellum brain samples (mean weight = 53.25 mg) were used for protein isolation. Samples were from a 59 yo female and 74 yo male, with PMIs of 30 and 22 hrs respectively, who had no known neurological or neuropsychiatric conditions. Samples were lysed in 500 μL of guanidinium-HCl buffer using tip-probe sonication, heated briefly to 95 ℃ and diluted 1:1 with LC-MS water before 4 mL of ice-cold acetone was added to precipitate protein overnight at -30 ℃. Following a wash (3 mL 80% cold acetone) and incubation (-30 ℃, 1 hr) supernatant was discarded and protein air-dried (RT, 30 mins). The protein pellet was resuspended in 500 μL 10% TFE in 100 mM HEPES (pH 7.5) and sonicated. Protein concentration was estimated with BCA (1 μL sample + 9 μL 2% SDS). Normalised protein (10 μg/10 μL) was then digested using a combination of LysC/trypsin or GluC for all samples. Peptides were separated on a Dionex 3500 nanoHPLC, coupled to an Orbitrap Lumos mass spectrometer (Thermo Fisher Scientific) via electrospray ionization in positive mode with 1.9 kV at 275 °C and RF set to 30%. Separation was achieved on a 50 cm × 75 µm column packed with C18AQ (1.9 µm; Dr Maisch, Ammerbuch, Germany) (PepSep, Marslev, Denmark) over 120 min at a flow rate of 300 nL/min. The peptides were eluted over a linear gradient of 3–40% Buffer B (Buffer A: 0.1% v/v formic acid; Buffer B: 80% v/v acetonitrile, 0.1% v/v FA) and the column was maintained at 50 °C. The instrument was operated in targeted M2 acquisition mode with an MS1 spectrum acquired over the mass range 300–1300 m/z (120,000 resolution, 100% automatic gain control (AGC) and 50 ms maximum injection time) followed by targeted MS/MS via HCD fragmentation with 0.7 m/z isolation (60,000 resolution, 200% AGC, 300 ms maximum injection time and stepped normalised collision energy 25, 30 and 35 eV). Data were analysed in Proteome Discover v2.5.0.400 with SequestHT [89] and searched against a custom .fasta database containing only risk gene predicted protein isoform sequences containing the targeted peptide sequences. Precursor mass tolerance was set to 10 ppm and fragment mass tolerance set to 0.02 Da. Data were filtered to 1% FDR at the peptide spectral match level only i.e. no protein level FDR, and MS/MS annotations were manually verified. Protein structure prediction was done using AlphaFold accessed through UCSF ChimeraX (v1.5) [45, 51, 90].

### Statistics

Statistical results presented in this manuscript included linear regression to investigate RNA yield and quality (RIN) against individual, pH and PMI. Ordinary one-way ANOVAs using Tukey’s multiple comparison correction were used to analyse isoform TPMs between brain regions. Statistical tests and associated graphical output were performed using GraphPad Prism 10.1.0.

### Isoform data visualisation

The publicly available web-tool IsoVis (v1.6, https://isomix.org/isovis/) was used to visualise RNA isoforms and associated expression data (Wan et al., 2024, *under review*). Known and novel RNA isoforms are represented as a stack to compare alternative splicing events between different isoforms. Read counts assigned to each RNA isoform for each of the 35 samples were visualised as a heatmap.

## Data Availability

Anonymised raw data will be uploaded to EGA prior to publication to allow access to approved users upon publication. All summarised data will be available upon publication or on request prior to publication.

## Declarations

### Ethics approval

Healthy control post-mortem human brain samples were obtained from six consented individuals collected by the Victorian Brain Bank (VBB) and the Human Research Ethics Committee of the University of Melbourne gave ethical approval for this work: #12457 and #28304.

### Consent for publication

The VBB obtained signed consent for whole-brain donation from either the donor or their next-of-kin in which the signed person states: “I agree that research data gathered from studies may be published providing the donor cannot be identified.” All samples mentioned in this study have been de-identified and, except VBB pathologist CM, authors were blinded to any other individual details beyond those mentioned in the methods.

### Availability of data and materials

All raw nanopore long-read data (fastq) generated for each of the genes reported in this manuscript are available at The European Genome-Phenome Archive (EGA) study: EGAS00001007744. Gene isoform GTFs and TPM data from IsoLamp analysis are available upon request. Scripts used for risk gene evidence curation and downstream data analysis are available upon request. IsoLamp is open source and freely available at https://github.com/ClarkLaboratory/IsoLamp. IsoVis is freely available at https://isomix.org/isovis/.

### Author contributions

MBC conceived the study. SJ, YP and JG wrote the IsoLamp software with testing assistance from RDP, RA, AL, EMW, YS, RK. SJ, RC and RDP wrote the code to analyse and collate the risk gene database. CM performed pathology analysis and classified post-mortem brain tissues. RDP performed RNA extraction, long-read experiments and bioinformatic analysis with assistance from YP, YY, RA, AL, AH, EMW, YS and RK. MD and BLP extracted protein, ran mass spectrophotometry and analysed results. MBC and RDP oversaw the research. RDP and MBC wrote the paper with input and review from all authors.

## Acknowledgements

The authors would like to acknowledge that brain tissues were received from the Victorian Brain Bank (VBB), supported by The Florey, The Alfred and the Victorian Institute of Forensic Medicine and funded in part by Parkinson’s Victoria, MND Victoria and FightMND. The authors would also like to thank Geoff Pavey at the VBB for his assistance with frozen tissue preparation. This research was supported by The University of Melbourne’s Research Computing Services and the Petascale Campus Initiative.

## Supplementary Figures

**Supplementary Figure 1.**
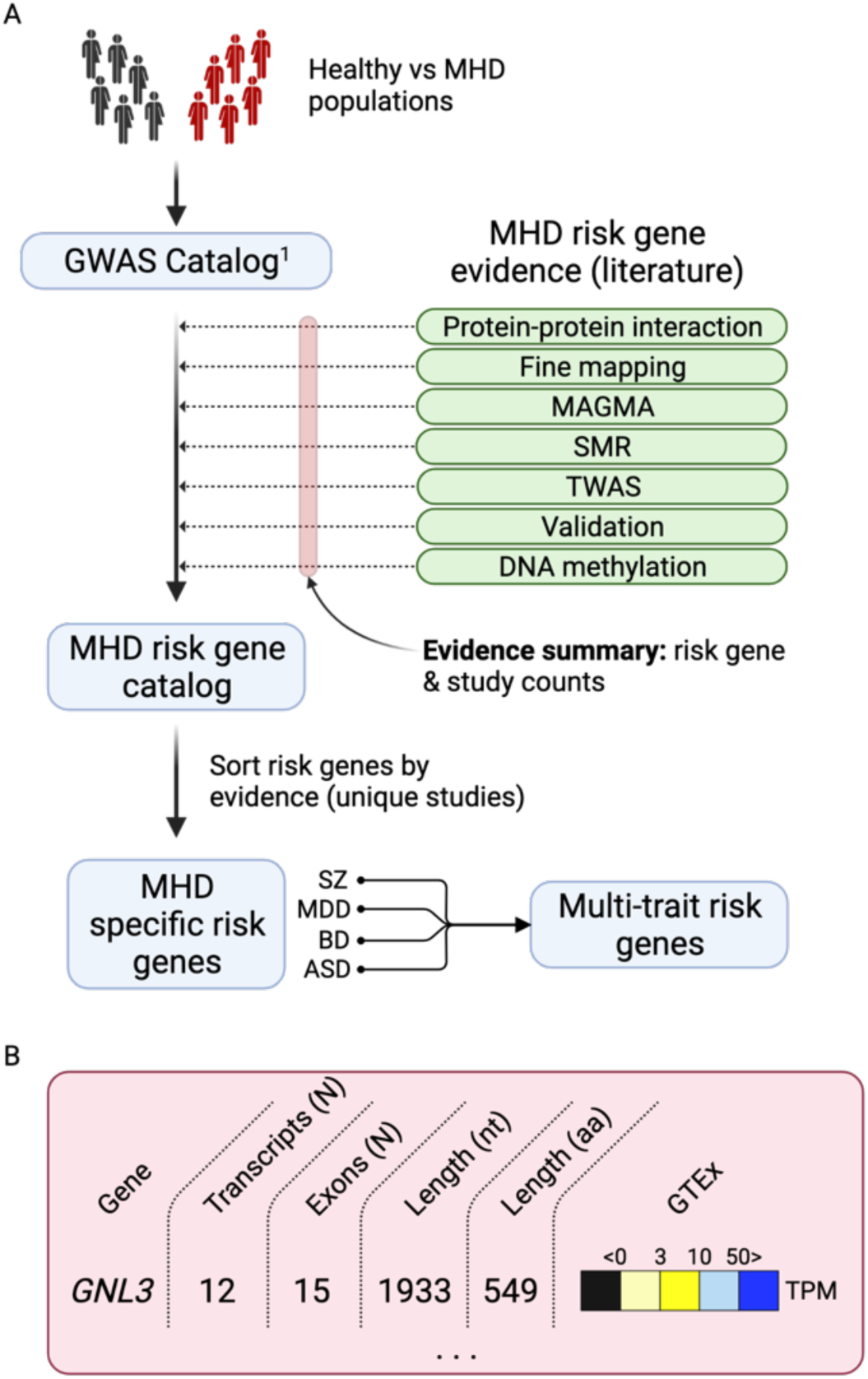
Mental health disorder (MHD) risk gene list curation pipeline. **A.** Single nucleotide polymorphism (SNP) catalogues form the foundation of the final risk gene evidence lists used to select genes for amplicon sequencing. These catalogues are collated from up-to-date, large-scale genome wide association studies (GWAS) of MHDs. A single GWAS catalogue was downloaded for schizophrenia (SZ), major depressive disorder (MDD), bipolar disorder (BD) and autism spectrum disorder (ASD), and associations were filtered according to criteria in Supplementary Table 4. GWAS data generally had either a mapped or reported gene associated with each SNP. Further evidence (i.e. reported genes) from categorised literature sources (shown in green) was then added to the list. This list was then sorted (high to low) by the number of occurrences of a risk gene in unique studies across all evidence/validation categories. No weighting was applied to any category. A multi-trait list was also made containing evidence for risk genes across all four MHDs so shared risk genes could be identified. **B.** An example of additional risk gene information included in the list e.g. for *GNL3*, count of known transcripts, count of coding exons for the canonical isoform, length in nucleotides (nt), length of protein in amino acids (aa) and the categorised Genotype-Tissue Expression (GTEx) in transcript per million (TPM) for each brain tissue [91]. *Definitions*: multi-marker analysis of Genomic annotation (MAGMA), summary-based Mendelian randomisation (SMR), transcriptome wide association study (TWAS).

**Supplementary Figure 2.**
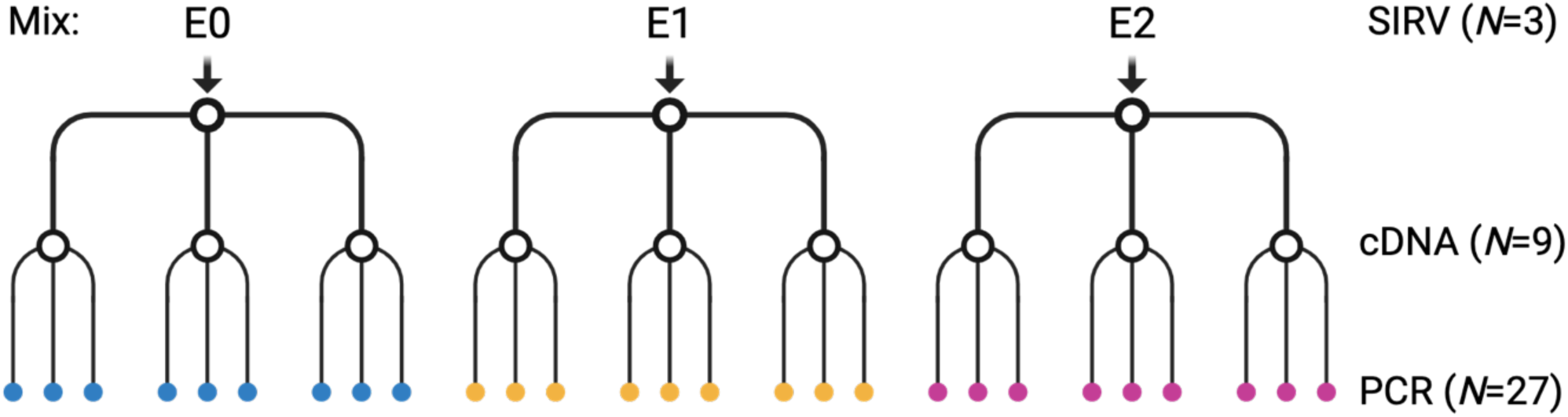
Experimental design of SIRV amplicon controls. E0, E1 and E2 represent each SIRV mix of known isoform concentrations. Each mix was converted into cDNA in triplicate and finally, the full length of synthetic genes SIRV5 and 6 were amplified using PCR in triplicate for each cDNA replicate.

**Supplementary Figure 3.**
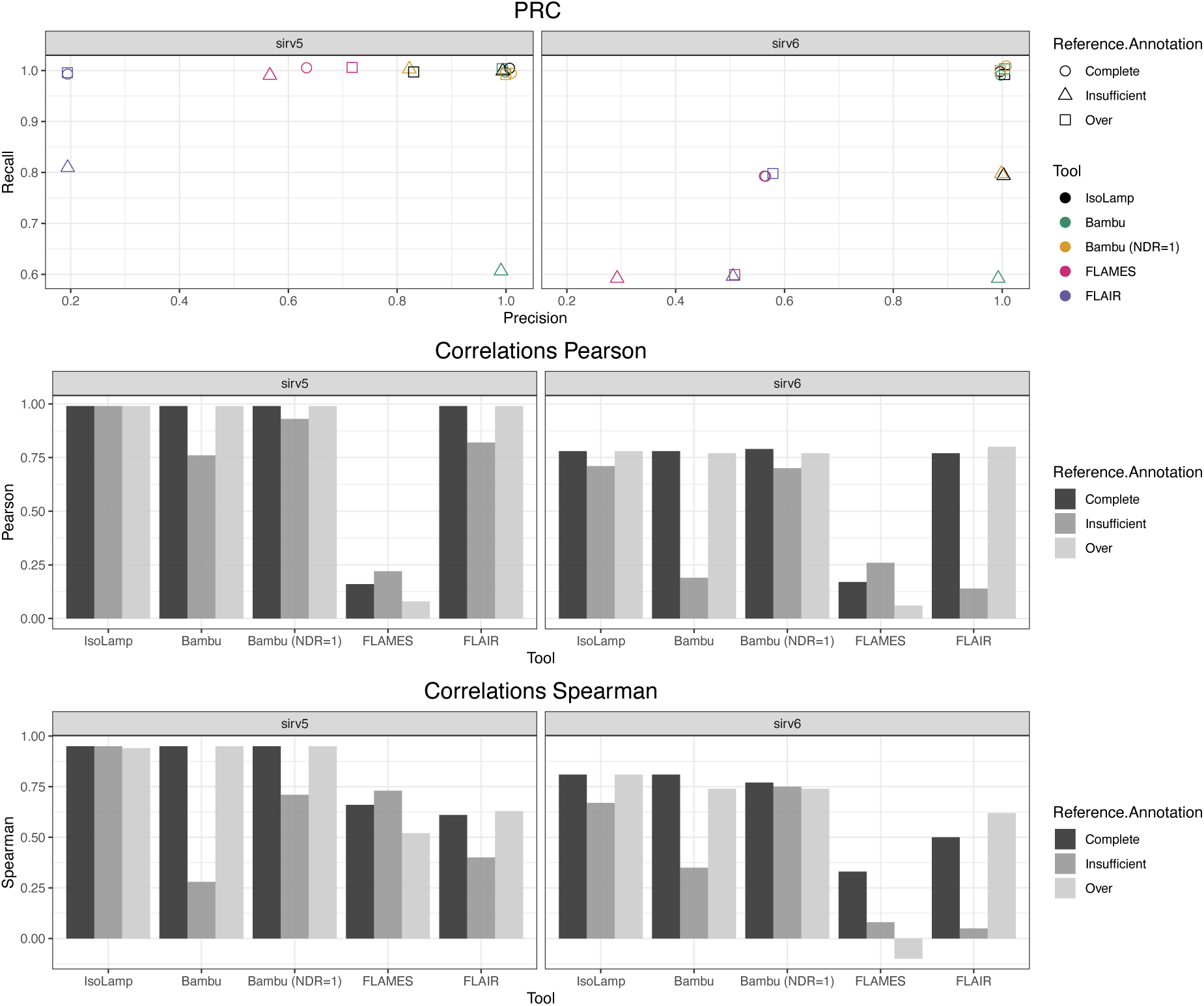
Benchmarking IsoLamp using spike-in SIRVs and the optimised IsoLamp expression-based filter. **A.** Precision recall of each tested pipeline with the complete, insufficient or over annotated SIRV reference, filtering all results using the IsoLamp expression-based filter. IsoLamp (black) returned high quality isoforms from amplicon data of both SIRV5 and 6. Pearson **(B)** and Spearman **(C)** correlations for each pipeline between known and observed expression values for SIRV 5 and 6 mixes.

**Supplementary Figure 4.**
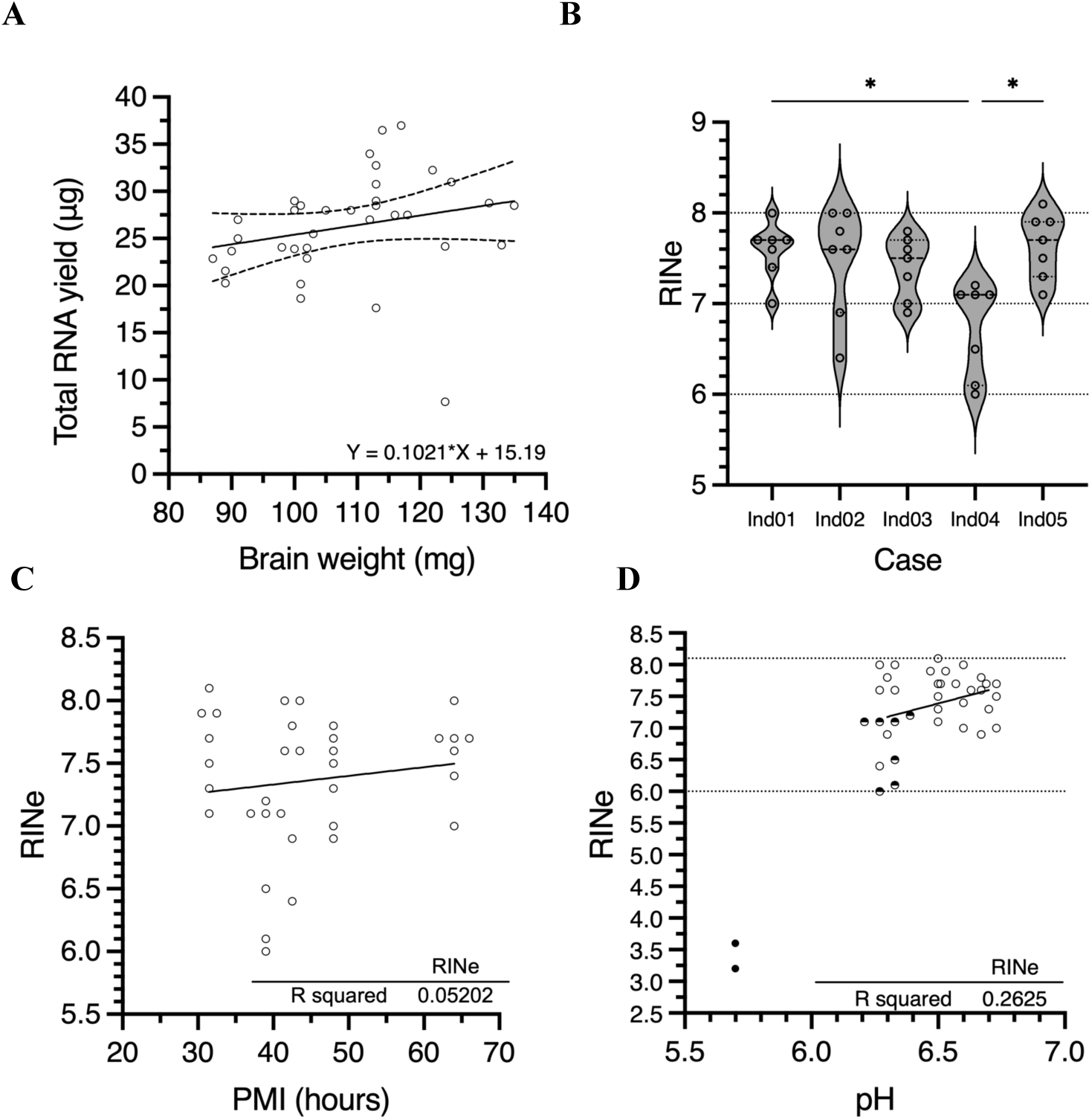
Post-mortem human brain RNA QC. **A.** Yield of isolated total RNA generally increases with increased tissue weight (mg) input to homogenisation using the RNeasy Lipid Tissue Mini Kit (QIAGEN:74804). **B.** The RNA integrity number equivalent (RINe) for brain tissue was generally between 7 – 8. RINe from individual (Ind) 04 was significantly lower when compared to individuals 01 (p=0.0259) and 05 (p=0.0433). One way ANOVA: F=6.224, DF=6. **C.** No correlation was detected between RNA quality (RINe) and individual post-mortem interval (PMI). **D.** Decreasing individual brain pH appears to impact RINe. Half-fill circles indicate samples from individual 04 (pH = 6.3). Filled circles indicate degraded RNA isolated from individual 06 (female)w which was not included for further analysis. Data in B, C and D are staggered for clarity.

**Supplementary Figure 5.**
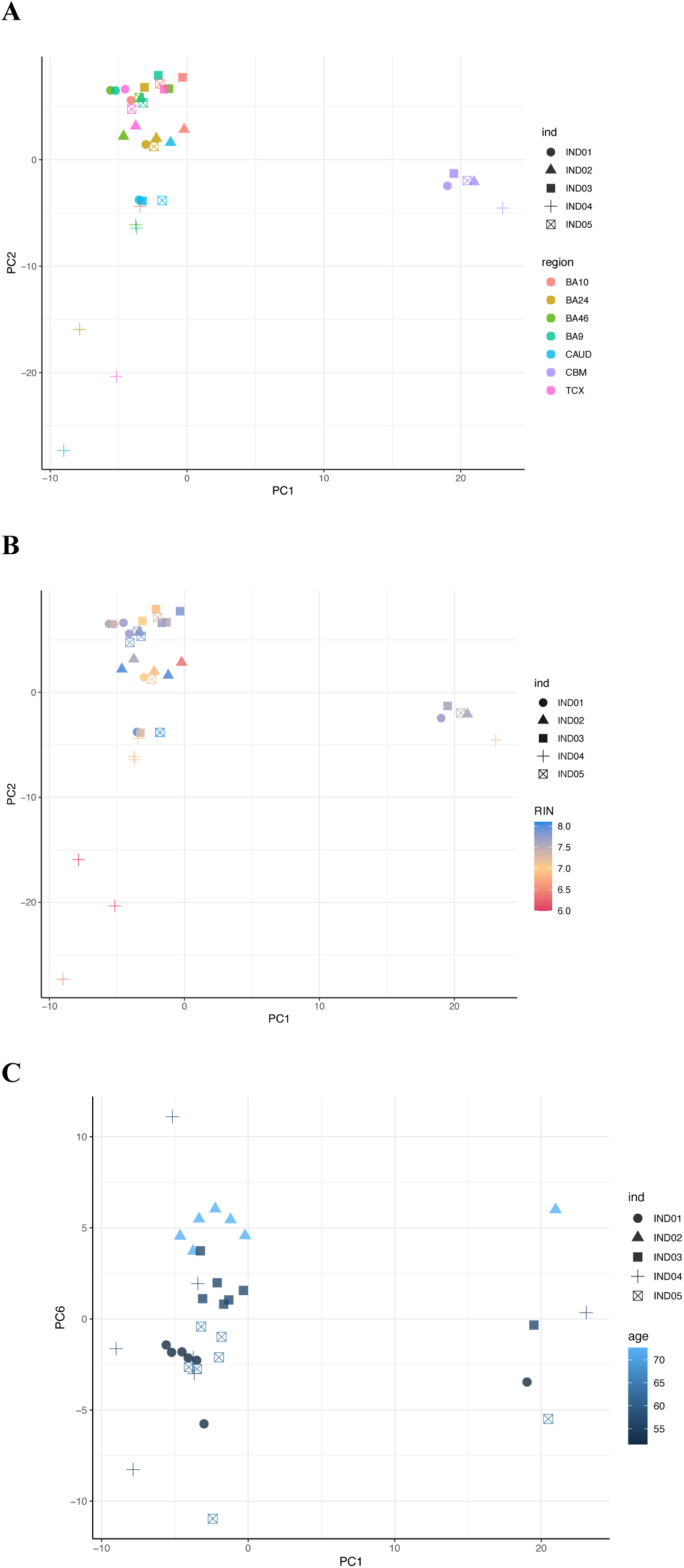
Principal component analyses (PCA) of brain samples. PC1 and PC2 coloured by brain region **(A)** and RNA integrity (RIN) **(B)**. **C.** PC4 and PC5 coloured by donor age (years). Key: individual (IND), Brodmann’s area (BA), caudate (caud), cerebellum (cbm) and temporal cortex (TCX).

**Supplementary Figure 6.**
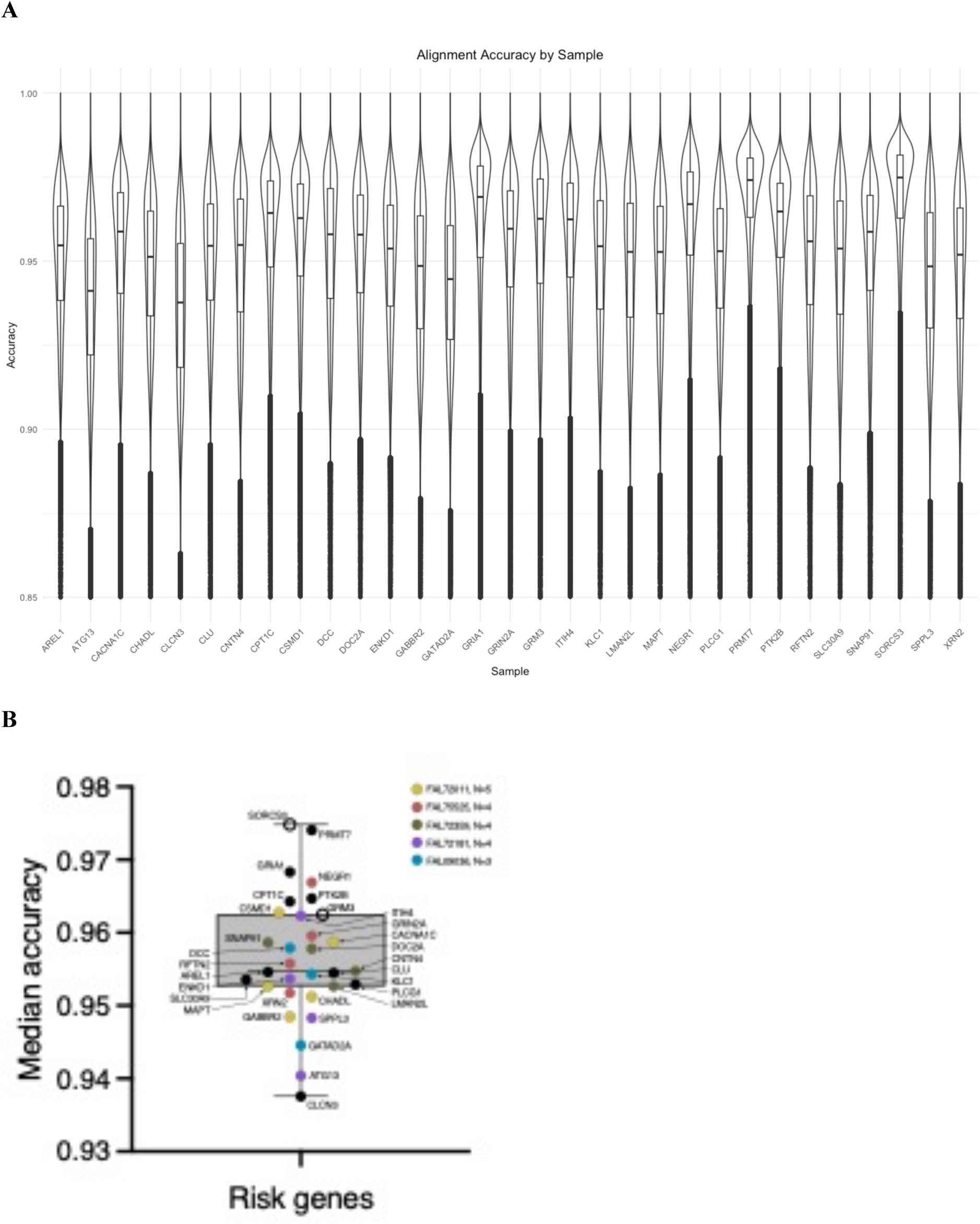
Long-read amplicon mapping accuracy. **A.** All sequenced risk genes. Plotted range 0.85 – 1.00. **B.** A box and whiskers plot of the median accuracy for each risk gene. Open circles indicate the library was prepared with ligation sequencing kit 110 (ONT). Colours indicate flow cells (prefix: FAL) that were used for multiple long-read libraries (N=3-5).

**Supplementary Figure 7.**
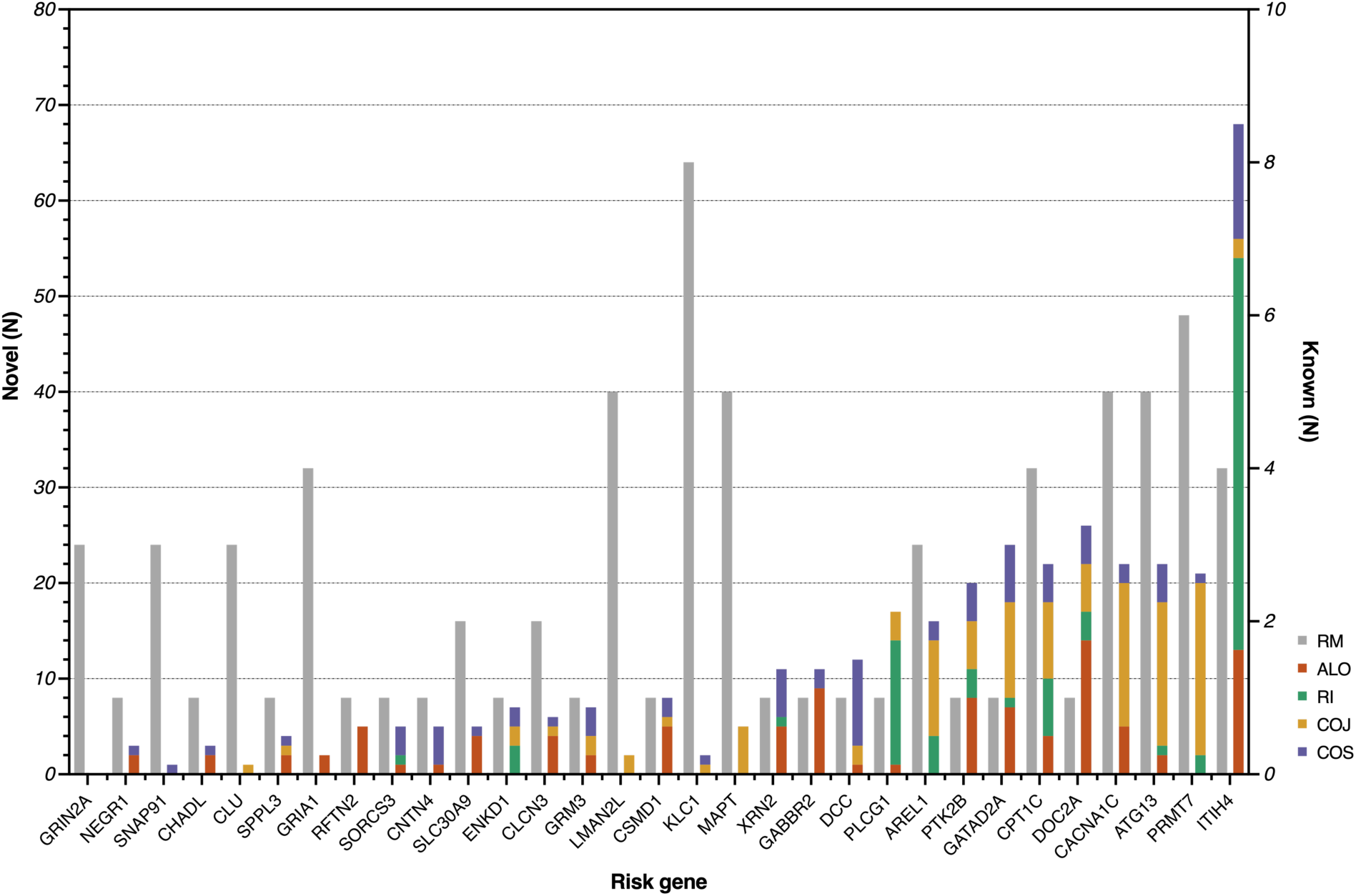
Risk gene isoform counts. The number of detected isoforms (known and novel) is shown for each risk gene sorted from lowest (*GRIN2A*) to highest (*ITIH4*). Each isoform was classified into a SQANTI subcategory: reference match (RM), containing at least one novel splice site (ALO), retained intron (RI), combination of known junctions (COJ) or splice sites (COS). Known (RM) isoform counts are plotted on the right Y-axis and novel isoform counts are plotted on the left Y-axis.

**Supplementary Figure 8.**
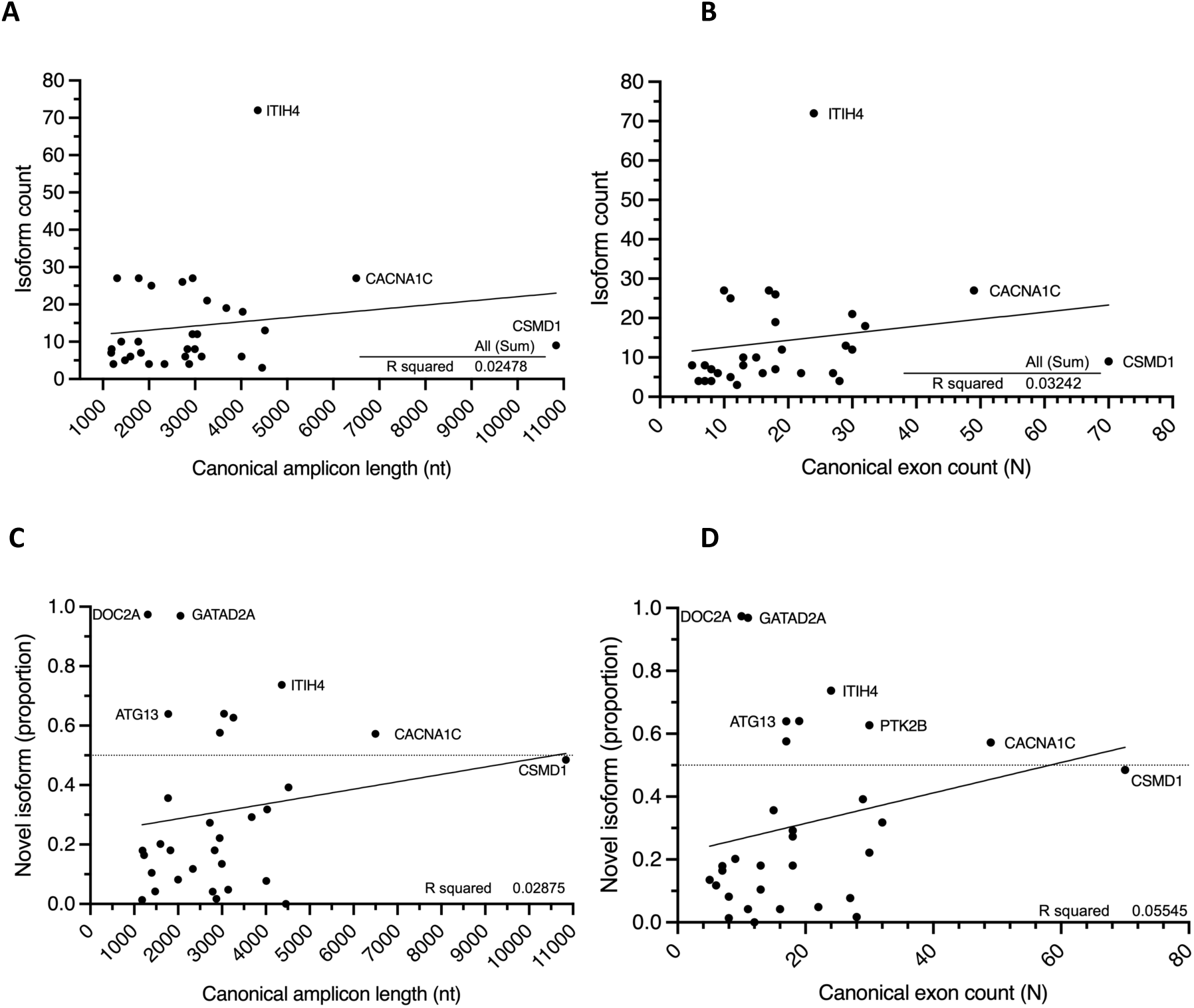
Linear regression of amplicon length or canonical exon count against isoform count and novel isoform TPM proportion does not deviate significantly from zero. Linear regression of known and novel isoform counts with expected canonical amplicon length (A) and number of canonical exons (B). Linear regression of novel isoform read proportion with expected canonical amplicon length (C) and number of canonical exons (D).

**Supplementary Figure 9.**
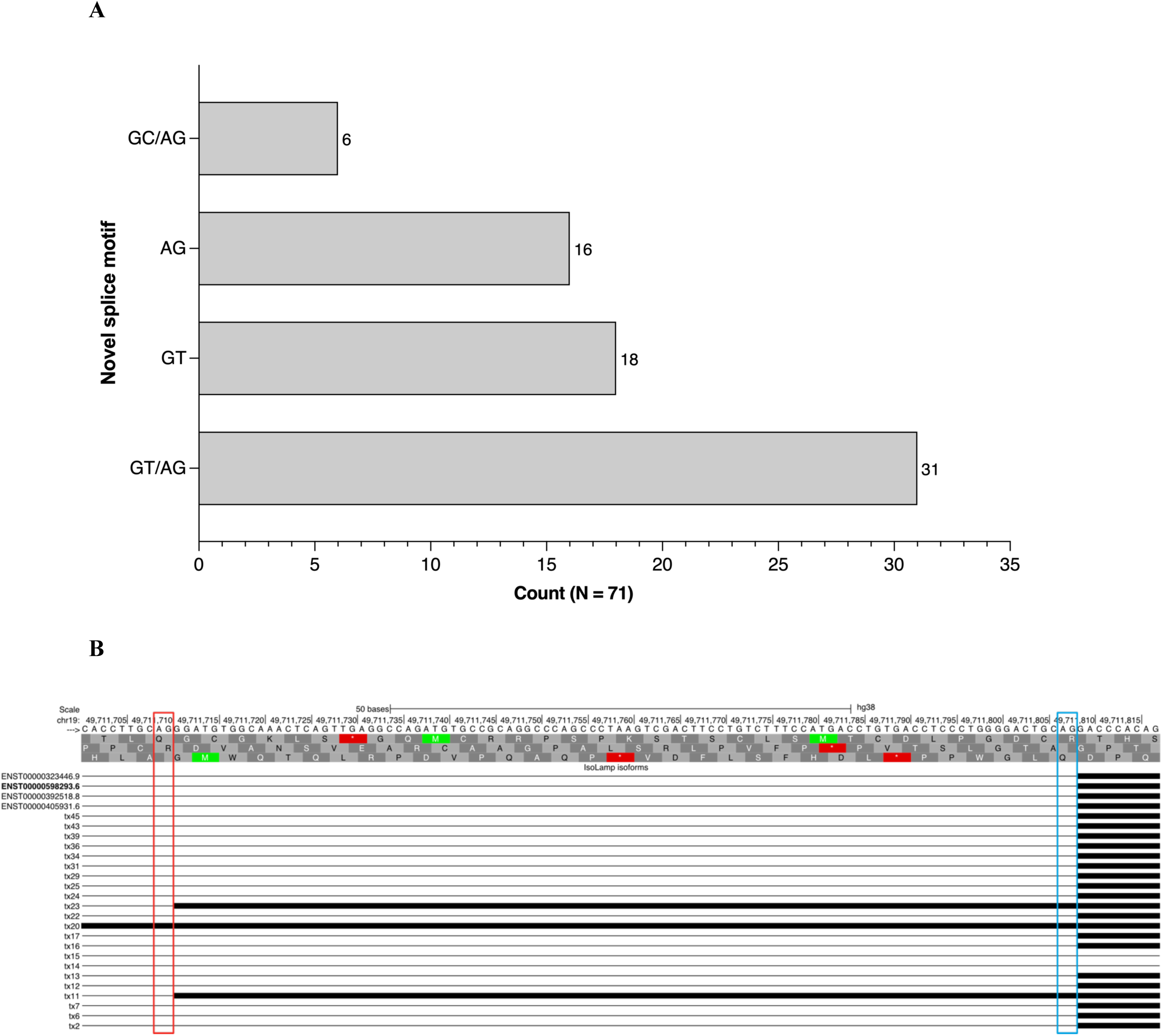
**A.** Count of novel isoform splice pairing. Isoforms classified as containing at least one novel splice site (ALO) were examined and the novel pair or donor/acceptor was counted. Duplicates were counted only once. **B.** UCSC screenshot of an example novel splice acceptor (red box, GT/**AG**, +98 nt) detected in two novel isoforms (Tx 11 and 23) in canonical exon 17 (blue box) for the schizophrenia risk gene *CPT1C*.

**Supplementary Figure 10.**
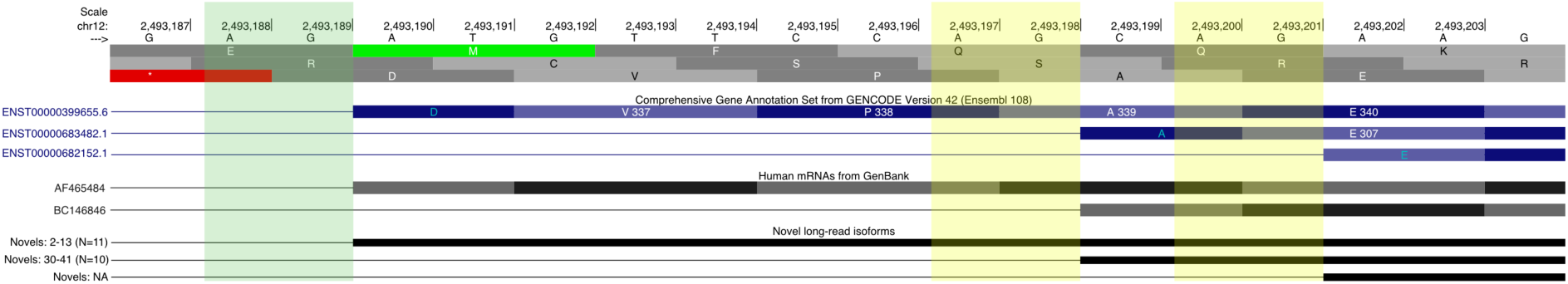
UCSC screenshot of *CACNA1C* splicing hotspot. Long-read sequencing identified 10 novel isoforms (black tracks) that support one of two annotated alternative splicing events (yellow boxes) within a 12 nt region (chr12:2,493,190-2,493,201) of exon 7 in *CACNA1C*. 11 novel isoforms also supported the use of the canonical (ENST00000399655.6) acceptor site (green box).

**Supplementary Figure 11.**
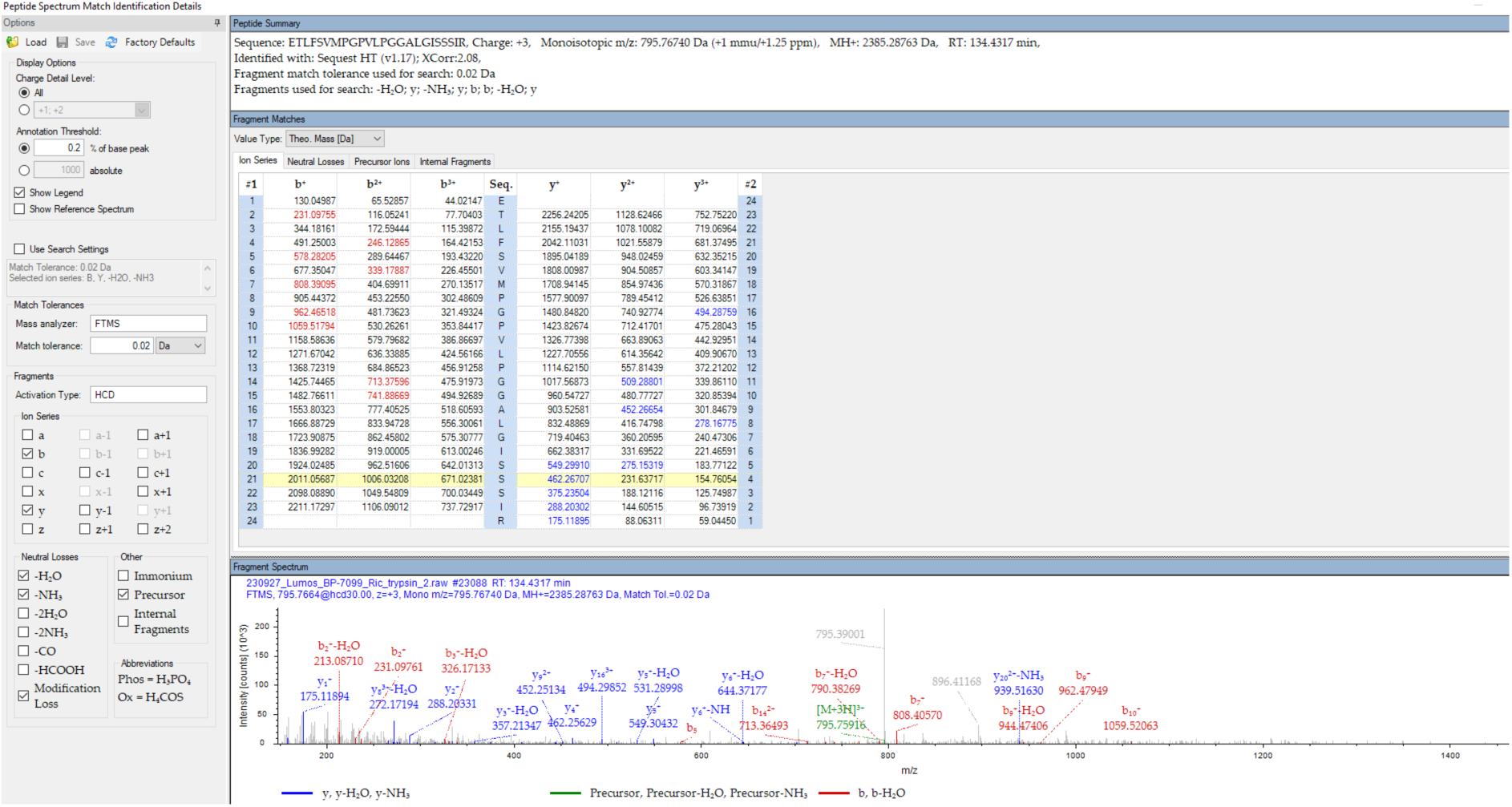
Annotated MS/MS spectrum highlighting matched b- and y-type ions providing proteomic [ETLFSVMPG//PVLPGGALGISSSIR] evidence for novel skipping of canonical exon 22 in *ITIH4* detected using long-reads.

**Supplementary Figure 12.**
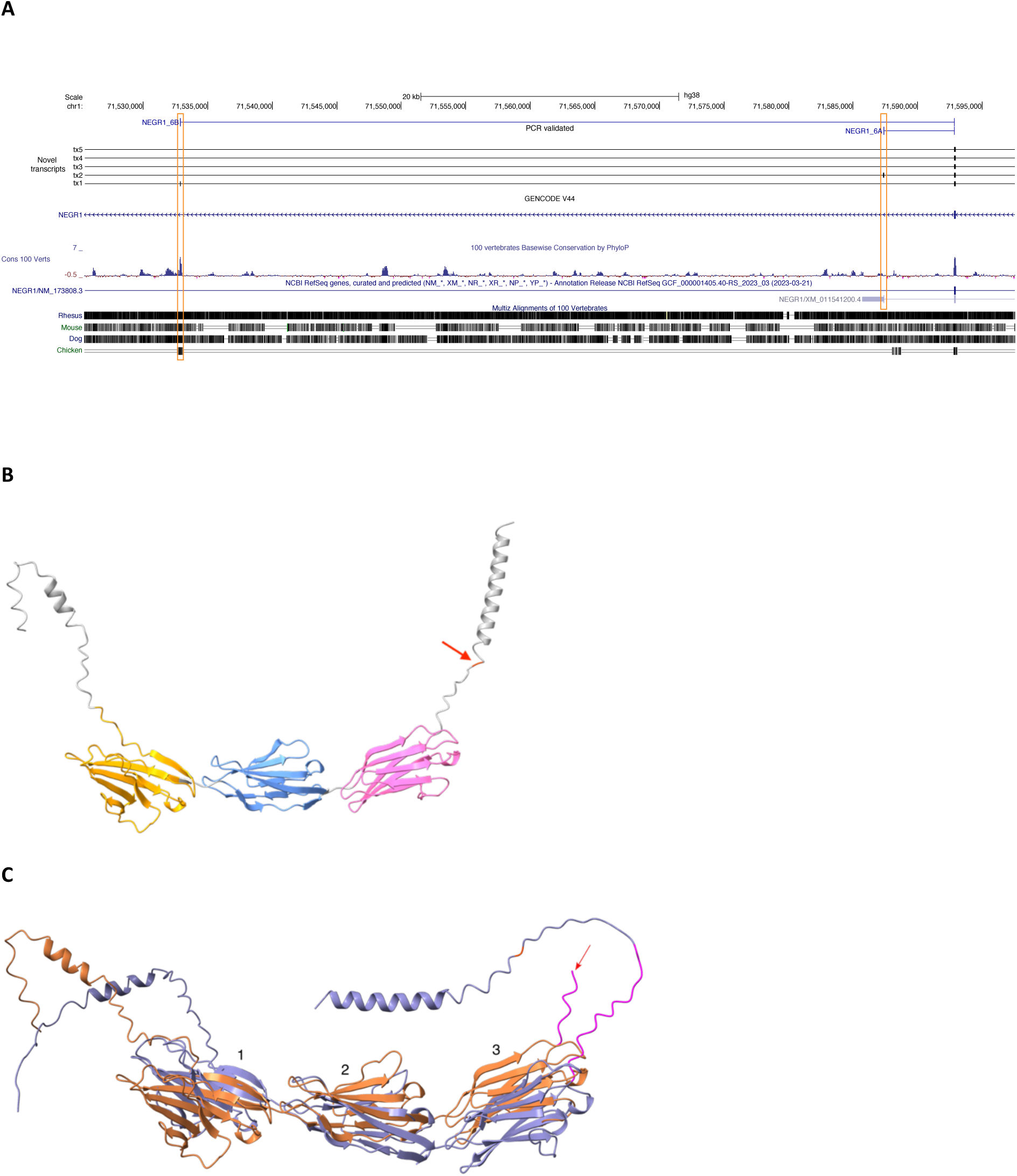
*NEGR1* splice isoforms and protein prediction. **A.** *NEGR1* novel exons were validated using Sanger sequencing of PCR amplicons and sequence reads were aligned and viewed using UCSC Genome Browser. Orange boxes indicate the novel exons and highlight high vertebrate conservation for exon 6b and predicted (NCBI RefSeq) termination site for exon 6a. **B.** AlphaFold protein prediction of the canonical *NEGR1* isoform (ENST00000357731). Three Ig-like domains are coloured according to the reported amino-acid positions (UniProt: Q7Z3B1); 1 (orange), 2 (blue) and 3 (pink). A GPI anchor residue (red) is shown at position 324 aa (C-terminal, red arrow). **C.** Overlaid AlphaFold protein predictions of novel Tx1 (purple) and Tx2 (orange). Ig-like domains are numbered 1-3, the GPI anchor (red) is present in Tx1 and the termination of Tx is indicated by a red arrow. Novel residues (pink) are indicated at the C-terminal end for both transcripts, Tx1: 14 aa and Tx2: 7 aa.

**Supplementary Figure 13.**
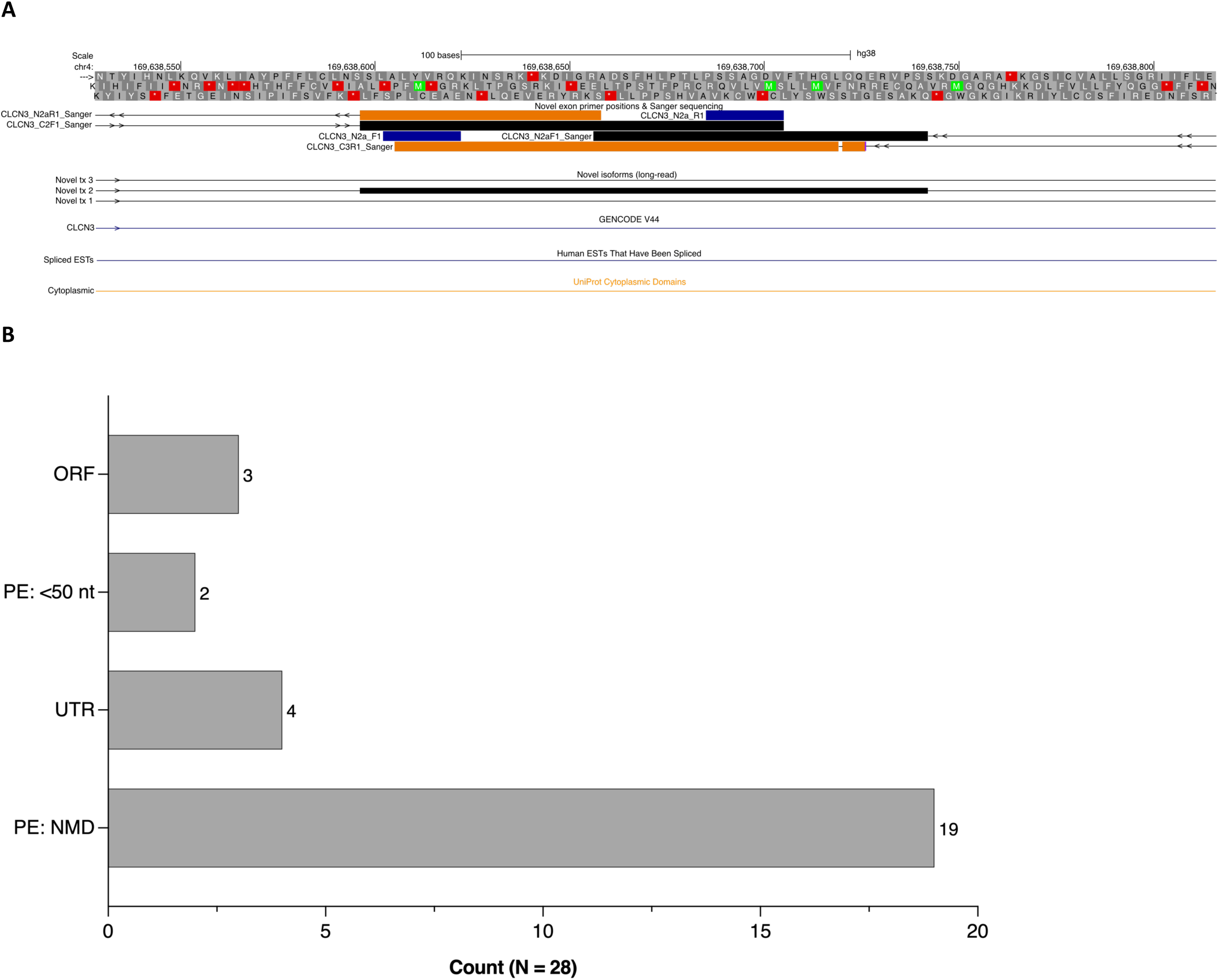
**A. Novel exon validation in *CLCN3*.** Novel exon 2a in the schizophrenia risk gene *CLCN3*, identified in long-read sequencing data, was validated by PCR using primers designed in the flanking cassette exons 1 and 3 (ENST00000513761). The novel exon sequence shown in novel transcript (Tx) 2 was validated with Sanger sequencing from the 5’ canonical exon 2 (C2F1) to the reverse primer within the novel sequence (N2aR1) and from the novel sequence (N2aF1) to the 3’ canonical exon 3 (C3R1). Direction of reads are indicated with arrows. Black and orange boxes indicate forward and reverse Sanger sequence respectively. Blue boxes indicate forward and reverse primers within the novel exon. A known cytoplasmic domain is shown by an orange track. Key: expressed sequence tag (EST). **B. Novel exon categories.** The impact of novel exon inclusion on the open reading frame of novel isoforms was predicted using Expasy [43] and then classed into groups, predicted to retain the open reading frame (ORF), inclusion of premature termination codon or ‘poison exon’ that was predicted to lead to nonsense-mediated decay (PE:NMD) or was <50 nt from the final exon junction (PE: <50 nt) or was with the 5’ or 3’ untranslated regions (UTR).

**Supplementary Figure 14.**
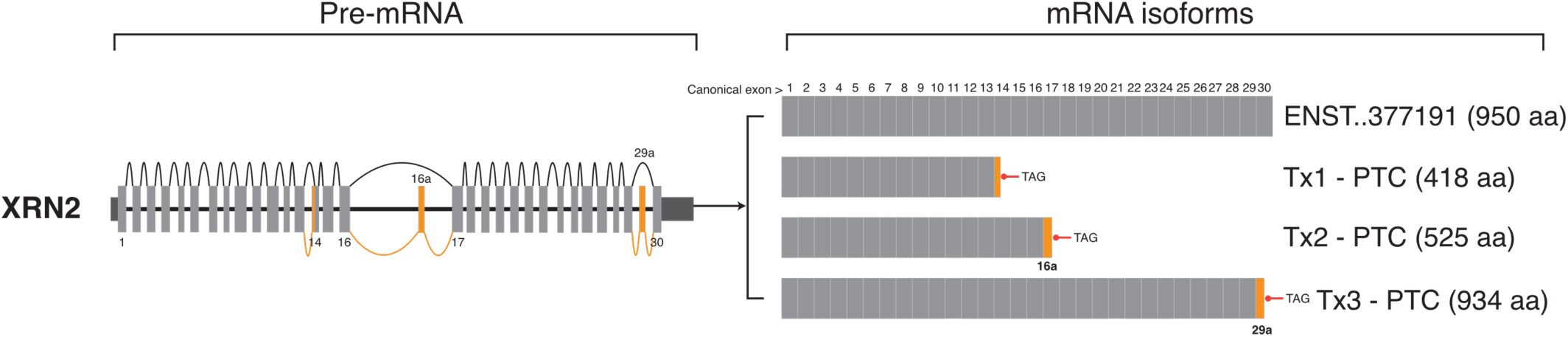
Splice graph of *XRN2* novel isoforms containing novel exons. Dark and light grey boxes indicate 5’ and 3’ UTR and coding exons respectively. Orange lines and boxes indicate novel splicing events and exons. Novel transcript 1 (Tx1) contains a novel splice acceptor (AG) within exon 14 (+24 nt) leading to a premature termination codon (PTC) and predicted nonsense mediated decay. Novel transcript 2 (Tx2) includes the validated novel exon 16a (54 nt) which was also predicted to encode a PTC. Novel isoform 3 (Tx3) contains a validated novel exon (29a) which encodes a PTC <50 nt from the final exon junction. Tx3 was predicted to lead to a truncated protein (934 aa). “..” indicates 0’s removed for brevity.

**Supplementary Figure 15.**
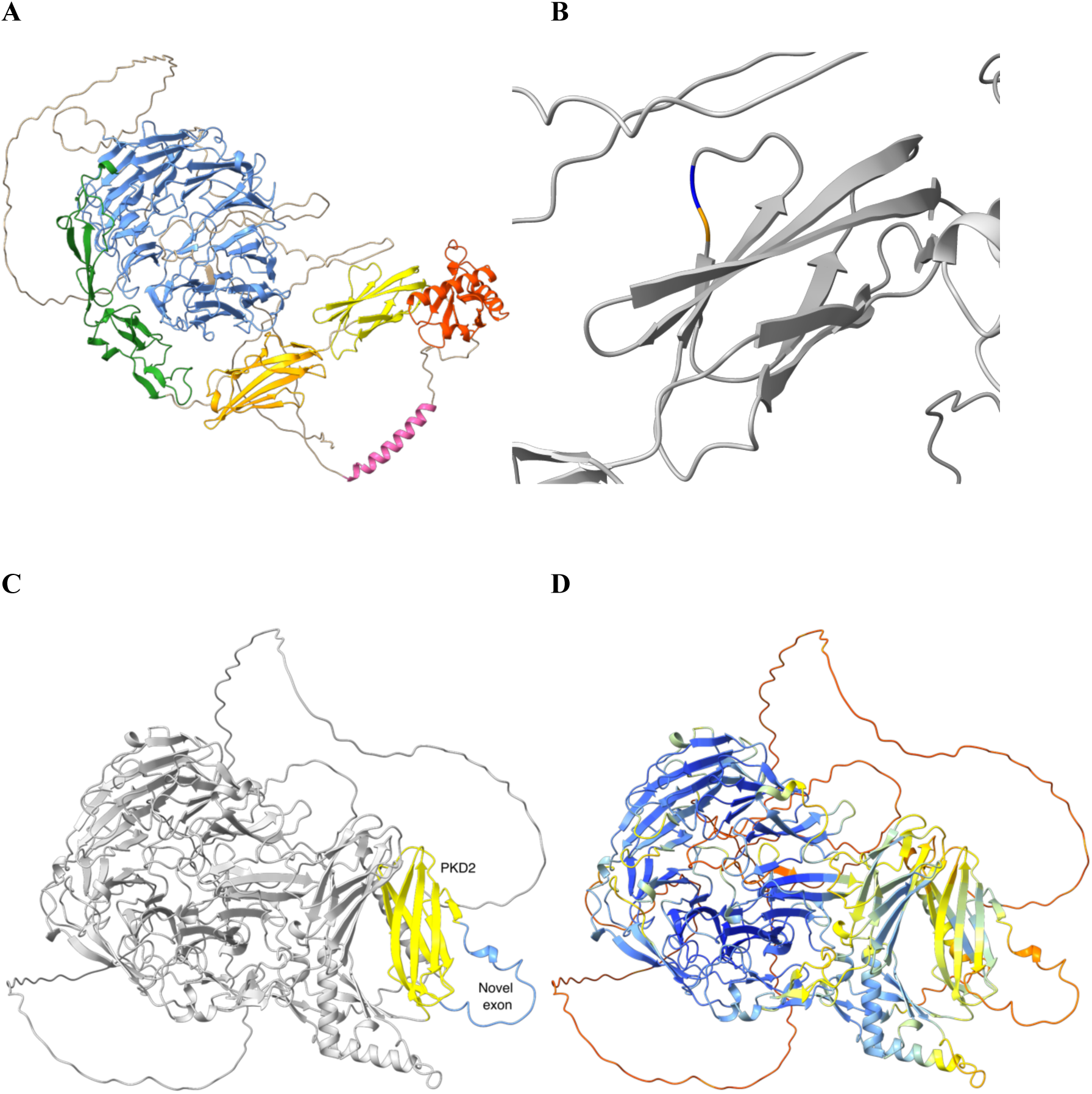
*SORCS3* novel exon and protein structure predictions. **A.** AlphaFold prediction of *SORCS3* canonical isoform (ENST00000369701.8: Q17R88) coloured by domain: β-propeller (blue), 10CC domain domains (green), polycystic kidney disease (PKD) domains PKD1 (orange) and PKD2 (yellow), SorCS membrane proximal (SoMP) (red) and transmembrane domain (pink). **B.** Zoomed view of the PKD2 domain indicating LYS:956 (blue) and PRO:957 (orange) where frame-retaining novel exon 20a (60 nt) is inserted. **C.** Protein structure prediction of novel transcript 1 (Tx1) containing the novel exon (blue) within the PKD2 domain (yellow). **D.** AlphaFold per-residue confidence scores (pLDDT) (0-100) for novel transcript 1: very high (>90, blue), confident (90-70, light-blue), low (70>50, yellow) and very low (<50, orange).

